## Appendix B for "COVID-19 vaccine uptake and effectiveness by time since vaccination in the Western Cape province, South Africa: An observational cohort study during 2020-2022"

This appendix provides supplementary analysis outputs for the following article:

2024

### Table of contents

**B1. Description of cohort 3**

B1.1 Cohort description by vaccination status 3

B1.2 Cohort description by age group 6

**B2. COVID-19 outcomes, SARS-CoV-2 testing and positivity, and COVID-19 vaccination over time 9**

B2.1 Outcomes, testing and vaccination over time: Males 10

B2.2 Outcomes, testing and vaccination over time: Females 11

B2.3 Outcomes, testing and vaccination over time: Age 18-34 years 12

B2.4 Outcomes, testing and vaccination over time: Age 35-49 years 13

B2.5 Outcomes, testing and vaccination over time: Age 50-59 years 14

B2.6 Outcomes, testing and vaccination over time: Age 60+ years 15

B2.7 Outcomes, testing and vaccination over time: Living with HIV at start 2020 16

B2.8 Outcomes, testing and vaccination over time: Not known to be living with HIV at start 2020 17

**B3. Vaccine effectiveness for distinct subpopulations 18**

B3.1 Vaccine effectiveness: Males 18

B3.2 Vaccine effectiveness: Females 19

B3.3 Vaccine effectiveness: Age 18-49 years 20

B3.4 Vaccine effectiveness: Age 50+ years 21

B3.5 Vaccine effectiveness: Known to be living with HIV at start 2020 22

B3.6 Vaccine effectiveness: Not known to be living with HIV at start 2020 23

B3.7 Vaccine effectiveness: Excluding people who received heterologous vaccines 24

**B4. All multivariable cox model hazard ratios 25**

B4.1 Hazard ratios: Demographic characteristics 25

B4.2 Hazard ratios: Non-communicable conditions 26

B4.3 Hazard ratios: Infectious conditions 26

B4.4 Hazard ratios: Other conditions 27

B4.5 Hazard ratios: Prior SARS-CoV-2 diagnosis 27

B4.6 Hazard ratios: Location 28

B4.7 Hazard ratios: Healthcare utilisation and testing 29

B4.8 Hazard ratios: Vaccine terms 30

**B5. Vaccine effectiveness for rolling windows of calendar time 31**

### B1. Description of cohort

The tables below report on all variables considered in this study. The cohort, which is followed during calendar years 2020 to 2022, is stratified by completion of a primary COVID-19 vaccination series by end 2022, or age group at start 2020. A subset of these variables is also reported in Tables 1 and 2 of the article. Percent (and count frequency) are reported.

#### *B1.1 Cohort description by vaccination status*

| | | Complete vaccination[a] by end 2022 | |
|---|---|---|---|
| | **Overall**<br>N = 2,429,927 | **No**<br>N = 1,455,385 | **Yes**<br>N = 974,542 |
| **Age category at start 2020 (years)** | | | |
| 18-34 | 41.6 (1,009,672) | 46.6 (677,813) | 34.1 (331,859) |
| 35-49 | 29.3 (711,077) | 27.2 (395,497) | 32.4 (315,580) |
| 50-59 | 14.1 (342,308) | 12.4 (179,961) | 16.7 (162,347) |
| 60-99 | 15.1 (366,870) | 13.9 (202,114) | 16.9 (164,756) |
| **Sex** | | | |
| Female | 59.7 (1,450,660) | 57.8 (841,578) | 62.5 (609,082) |
| Male | 40.3 (979,267) | 42.2 (613,807) | 37.5 (365,460) |
| **PHC visit location type** | | | |
| Cape Town Metro | 51.4 (1,248,277) | 51.0 (742,774) | 51.9 (505,503) |
| Other (non-Metro or no visit recorded) | 48.6 (1,181,650) | 49.0 (712,611) | 48.1 (469,039) |
| **Location (subdistrict or district)** | | | |
| S01 | 7.3 (176,386) | 7.4 (107,731) | 7.0 (68,655) |
| S02 | 3.9 (94,234) | 3.9 (57,051) | 3.8 (37,183) |
| S03 | 6.1 (147,607) | 5.6 (81,768) | 6.8 (65,839) |
| S04 | 9.4 (227,820) | 9.1 (132,798) | 9.8 (95,022) |
| S05 | 13.4 (325,336) | 12.6 (183,230) | 14.6 (142,106) |
| S06 | 1.4 (35,067) | 1.6 (23,180) | 1.2 (11,887) |
| S07 | 11.4 (276,582) | 12.5 (181,870) | 9.7 (94,712) |
| S08 | 8.3 (200,836) | 8.3 (121,121) | 8.2 (79,715) |
| S09 | 6.3 (153,580) | 6.4 (92,565) | 6.3 (61,015) |
| S10 | 8.9 (215,155) | 9.0 (130,917) | 8.6 (84,238) |
| S11 | 5.3 (127,820) | 5.1 (73,827) | 5.5 (53,993) |
| S12 | 11.3 (273,812) | 11.2 (162,500) | 11.4 (111,312) |
| S13 | 7.2 (175,692) | 7.3 (106,827) | 7.1 (68,865) |

| | Overall N = 2,429,927 | Complete vaccination[a] by end 2022 No N = 1,455,385 | Yes N = 974,542 |
|---|---|---|---|
| **Number of PHC facility visits (during 5 years before the pandemic)** | | | |
| 0 | 17.2 (418,046) | 17.8 (258,816) | 16.3 (159,230) |
| 1-4 | 31.3 (759,411) | 33.8 (492,631) | 27.4 (266,780) |
| 5-14 | 23.4 (567,415) | 23.7 (345,062) | 22.8 (222,353) |
| 15+ | 28.2 (685,055) | 24.7 (358,876) | 33.5 (326,179) |
| **Number of years with PHC facility visits (out of the 3 years before the pandemic)** | | | |
| 0 | 26.9 (654,432) | 28.9 (420,879) | 24.0 (233,553) |
| 1 | 24.5 (594,713) | 26.4 (384,147) | 21.6 (210,566) |
| 2 | 18.1 (440,456) | 18.6 (270,732) | 17.4 (169,724) |
| 3 | 30.5 (740,326) | 26.1 (379,627) | 37.0 (360,699) |
| **Hypertension** | | | |
| Absent by end 2022 | 76.5 (1,858,079) | 79.8 (1,161,884) | 71.4 (696,195) |
| Onset during 2020-2022 | 3.4 (83,174) | 2.8 (40,473) | 4.4 (42,701) |
| Present before 2020 | 20.1 (488,674) | 17.4 (253,028) | 24.2 (235,646) |
| **Diabetes** | | | |
| Absent by end 2022 | 89.5 (2,174,691) | 90.8 (1,321,701) | 87.5 (852,990) |
| Onset during 2020-2022 | 1.9 (46,111) | 1.6 (22,634) | 2.4 (23,477) |
| Present before 2020 | 8.6 (209,125) | 7.6 (111,050) | 10.1 (98,075) |
| **Chronic kidney disease** | | | |
| Absent by end 2022 | 95.7 (2,324,768) | 95.9 (1,396,419) | 95.3 (928,349) |
| Onset during 2020-2022 | 0.7 (16,445) | 0.5 (7,707) | 0.9 (8,738) |
| Present before 2020 | 3.7 (88,714) | 3.5 (51,259) | 3.8 (37,455) |
| **Chronic respiratory disease** | | | |
| Absent by end 2022 | 92.2 (2,241,094) | 92.5 (1,346,451) | 91.8 (894,643) |
| Onset during 2020-2022 | 1.5 (36,451) | 1.4 (20,454) | 1.6 (15,997) |
| Present before 2020 | 6.3 (152,382) | 6.1 (88,480) | 6.6 (63,902) |
| **HIV** | | | |
| Absent by end 2022 | 85.4 (2,075,796) | 85.3 (1,241,925) | 85.6 (833,871) |
| Onset during 2020-2022 | 1.4 (32,986) | 1.4 (21,048) | 1.2 (11,938) |
| Present before 2020 | 13.2 (321,145) | 13.2 (192,412) | 13.2 (128,733) |
| **Tuberculosis (at any time in past)[b]** | | | |
| Absent by end 2022 | 90.2 (2,190,920) | 89.2 (1,298,552) | 91.6 (892,368) |
| (First) onset during 2020-2022 | 1.7 (42,068) | 2.0 (28,783) | 1.4 (13,285) |
| Present before 2020 | 8.1 (196,939) | 8.8 (128,050) | 7.1 (68,889) |
| **Tuberculosis (ongoing episode)** | | | |
| Absent throughout 2020-2022 | 96.8 (2,351,220) | 96.2 (1,400,628) | 97.5 (950,592) |
| Experienced episode during 2020-2022 | 3.2 (78,707) | 3.8 (54,757) | 2.5 (23,950) |
| **Pregnancy (ongoing episode)** | | | |
| Absent throughout 2020-2022 | 91.5 (2,222,462) | 90.6 (1,319,172) | 92.7 (903,290) |
| Experienced during 2020-2022 | 8.5 (207,465) | 9.4 (136,213) | 7.3 (71,252) |

| | Overall<br>N = 2,429,927 | Complete vaccination[a] by end 2022<br>No<br>N = 1,455,385 | <br>Yes<br>N = 974,542 |
|---|---|---|---|
| **Number of negative SARS-CoV-2 tests (by end 2022)** | | | |
| 0 | 81.8 (1,987,267) | 84.3 (1,227,129) | 78.0 (760,138) |
| 1 | 13.1 (317,758) | 11.4 (165,723) | 15.6 (152,035) |
| 2+ | 5.1 (124,902) | 4.3 (62,533) | 6.4 (62,369) |
| **COVID-19 vaccination status[a] and boosters (at end 2022)** | | | |
| None | 55.0 (1,336,826) | 91.9 (1,336,826) | 0.0 (0) |
| Incomplete BNT162b2 | 4.9 (118,559) | 8.1 (118,559) | 0.0 (0) |
| Complete Ad26.COV2S (no booster) | 9.7 (236,725) | 0.0 (0) | 24.3 (236,725) |
| Complete Ad26.COV2S (and booster) | 1.7 (41,196) | 0.0 (0) | 4.2 (41,196) |
| Complete BNT162b2 (no booster) | 22.6 (548,914) | 0.0 (0) | 56.3 (548,914) |
| Complete BNT162b2 (and booster) | 6.1 (147,707) | 0.0 (0) | 15.2 (147,707) |
| **Vital status (at end 2022)** | | | |
| Alive | 95.4 (2,318,859) | 93.3 (1,357,339) | 98.7 (961,520) |
| Died: not COVID-19 related | 4.1 (99,525) | 6.0 (87,001) | 1.3 (12,524) |
| Died: COVID-19 related | 0.5 (11,543) | 0.8 (11,045) | 0.1 (498) |
| **SARS-CoV-2 diagnosis (by end 2022)** | | | |
| Not experienced | 92.1 (2,237,396) | 93.3 (1,357,422) | 90.3 (879,974) |
| Experienced (2020-2022) | 7.9 (192,531) | 6.7 (97,963) | 9.7 (94,568) |
| **COVID-19 related hospitalisation or death (by end 2022)** | | | |
| Not experienced | 98.4 (2,391,586) | 98.2 (1,429,692) | 98.7 (961,894) |
| Experienced (2020-2022) | 1.6 (38,341) | 1.8 (25,693) | 1.3 (12,648) |
| **COVID-19 related hospitalisation with severe disease or death (by end 2022)** | | | |
| Not experienced | 99.3 (2,412,279) | 99.0 (1,441,138) | 99.7 (971,141) |
| Experienced (2020-2022) | 0.7 (17,648) | 1.0 (14,247) | 0.3 (3,401) |
| **COVID-19 related death (by end 2022)** | | | |
| Not experienced | 99.5 (2,418,384) | 99.2 (1,444,340) | 99.9 (974,044) |
| Experienced (2020-2022) | 0.5 (11,543) | 0.8 (11,045) | 0.1 (498) |

PHC: Primary healthcare
[a] Complete vaccination: ≥28 days since a first Ad26.COV2.S dose or ≥14 days since a second dose following a first BNT162b2 dose; incomplete BNT162b2: ≥21 days since a first BNT162b2 dose but not yet completely vaccinated (see methods).
[b] Tuberculosis (at any time in past) includes those with an ongoing episode.

#### B1.2 Cohort description by age group

| Characteristic | Overall | Age category (years) at start 2020 | | | |
|---|---|---|---|---|---|
| | | 18-34 | 35-49 | 50-59 | 60-99 |
| | N = 2,429,927 | N = 1,009,672 | N = 711,077 | N = 342,308 | N = 366,870 |
| **Sex** | | | | | |
| Female | 59.7 (1,450,660) | 62.0 (626,011) | 57.6 (409,758) | 56.3 (192,795) | 60.5 (222,096) |
| Male | 40.3 (979,267) | 38.0 (383,661) | 42.4 (301,319) | 43.7 (149,513) | 39.5 (144,774) |
| **PHC visit location type** | | | | | |
| Cape Town Metro | 51.4 (1,248,277) | 52.5 (530,567) | 51.2 (363,887) | 48.8 (167,199) | 50.9 (186,624) |
| Other (non-Metro or no visit recorded) | 48.6 (1,181,650) | 47.5 (479,105) | 48.8 (347,190) | 51.2 (175,109) | 49.1 (180,246) |
| **Location (subdistrict or district)** | | | | | |
| S01 | 7.3 (176,386) | 7.7 (77,751) | 7.4 (52,369) | 6.8 (23,243) | 6.3 (23,023) |
| S02 | 3.9 (94,234) | 4.2 (42,196) | 4.0 (28,349) | 3.2 (11,123) | 3.4 (12,566) |
| S03 | 6.1 (147,607) | 5.2 (52,567) | 5.7 (40,740) | 6.7 (22,812) | 8.6 (31,488) |
| S04 | 9.4 (227,820) | 8.7 (87,823) | 9.5 (67,544) | 9.6 (32,846) | 10.8 (39,607) |
| S05 | 13.4 (325,336) | 13.5 (136,218) | 13.3 (94,447) | 14.2 (48,470) | 12.6 (46,201) |
| S06 | 1.4 (35,067) | 1.3 (13,396) | 1.5 (10,403) | 1.6 (5,604) | 1.5 (5,664) |
| S07 | 11.4 (276,582) | 11.0 (110,967) | 11.5 (81,646) | 12.1 (41,279) | 11.6 (42,690) |
| S08 | 8.3 (200,836) | 9.8 (99,447) | 8.7 (62,133) | 6.3 (21,431) | 4.9 (17,825) |
| S09 | 6.3 (153,580) | 6.2 (62,751) | 6.2 (43,887) | 6.3 (21,576) | 6.9 (25,366) |
| S10 | 8.9 (215,155) | 9.5 (96,164) | 9.0 (63,647) | 7.6 (26,156) | 8.0 (29,188) |
| S11 | 5.3 (127,820) | 5.4 (54,433) | 5.2 (37,277) | 5.1 (17,404) | 5.1 (18,706) |
| S12 | 11.3 (273,812) | 10.1 (102,187) | 11.0 (78,196) | 12.9 (44,020) | 13.5 (49,409) |
| S13 | 7.2 (175,692) | 7.3 (73,772) | 7.1 (50,439) | 7.7 (26,344) | 6.9 (25,137) |

| Characteristic | Overall N = 2,429,927 | Age category (years) at start 2020: 18-34 N = 1,009,672 | 35-49 N = 711,077 | 50-59 N = 342,308 | 60-99 N = 366,870 |
|---|---|---|---|---|---|
| **Number of PHC facility visits (during 5 years before the pandemic)** | | | | | |
| 0 | 17.2 (418,046) | 16.5 (166,823) | 17.8 (126,341) | 17.5 (59,992) | 17.7 (64,890) |
| 1-4 | 31.3 (759,411) | 37.7 (380,587) | 31.4 (223,158) | 24.8 (84,912) | 19.3 (70,754) |
| 5-14 | 23.4 (567,415) | 27.4 (276,607) | 21.7 (154,654) | 19.9 (68,030) | 18.6 (68,124) |
| 15+ | 28.2 (685,055) | 18.4 (185,655) | 29.1 (206,924) | 37.8 (129,374) | 44.5 (163,102) |
| **Number of years with PHC facility visits (out of the 3 years before the pandemic)** | | | | | |
| 0 | 26.9 (654,432) | 26.4 (266,477) | 28.7 (203,854) | 26.3 (90,179) | 25.6 (93,922) |
| 1 | 24.5 (594,713) | 29.1 (294,248) | 24.1 (171,661) | 19.9 (68,264) | 16.5 (60,540) |
| 2 | 18.1 (440,456) | 22.0 (221,637) | 17.1 (121,763) | 14.9 (50,967) | 12.6 (46,089) |
| 3 | 30.5 (740,326) | 22.5 (227,310) | 30.1 (213,799) | 38.8 (132,898) | 45.3 (166,319) |
| **Hypertension** | | | | | |
| Absent at end 2022 | 76.5 (1,858,079) | 94.8 (957,219) | 79.2 (563,427) | 55.5 (190,033) | 40.2 (147,400) |
| Onset during 2020-2022 | 3.4 (83,174) | 1.7 (17,495) | 4.4 (31,142) | 5.6 (19,083) | 4.2 (15,454) |
| Present before 2020 | 20.1 (488,674) | 3.5 (34,958) | 16.4 (116,508) | 38.9 (133,192) | 55.6 (204,016) |
| **Diabetes** | | | | | |
| Absent at end 2022 | 89.5 (2,174,691) | 97.8 (987,299) | 91.8 (652,528) | 80.3 (274,806) | 70.9 (260,058) |
| Onset during 2020-2022 | 1.9 (46,111) | 0.7 (7,051) | 1.9 (13,643) | 3.6 (12,180) | 3.6 (13,237) |
| Present before 2020 | 8.6 (209,125) | 1.5 (15,322) | 6.3 (44,906) | 16.2 (55,322) | 25.5 (93,575) |
| **Chronic kidney disease** | | | | | |
| Absent at end 2022 | 95.7 (2,324,768) | 99.6 (1,005,786) | 98.1 (697,237) | 94.6 (323,845) | 81.2 (297,900) |
| Onset during 2020-2022 | 0.7 (16,445) | 0.1 (1,061) | 0.5 (3,576) | 1.1 (3,866) | 2.2 (7,942) |
| Present before 2020 | 3.7 (88,714) | 0.3 (2,825) | 1.4 (10,264) | 4.3 (14,597) | 16.6 (61,028) |
| **Chronic respiratory disease** | | | | | |
| Absent at end 2022 | 92.2 (2,241,094) | 97.1 (980,010) | 93.7 (665,959) | 86.2 (294,983) | 81.8 (300,142) |
| Onset during 2020-2022 | 1.5 (36,451) | 0.7 (7,179) | 1.4 (10,079) | 2.5 (8,717) | 2.9 (10,476) |
| Present before 2020 | 6.3 (152,382) | 2.2 (22,483) | 4.9 (35,039) | 11.3 (38,608) | 15.3 (56,252) |
| **HIV** | | | | | |
| Absent at end 2022 | 85.4 (2,075,796) | 85.2 (860,276) | 77.9 (553,891) | 89.4 (305,946) | 97.0 (355,683) |
| Onset during 2020-2022 | 1.4 (32,986) | 2.1 (21,170) | 1.3 (8,973) | 0.6 (2,127) | 0.2 (716) |
| Present before 2020 | 13.2 (321,145) | 12.7 (128,226) | 20.8 (148,213) | 10.0 (34,235) | 2.9 (10,471) |
| **Tuberculosis (at any time in past)[a]** | | | | | |
| Absent at end 2022 | 90.2 (2,190,920) | 91.5 (923,917) | 86.9 (617,997) | 88.8 (303,895) | 94.1 (345,111) |
| (First) onset during 2020-2022 | 1.7 (42,068) | 1.9 (19,093) | 1.9 (13,683) | 1.6 (5,604) | 1.0 (3,688) |
| Present before 2020 | 8.1 (196,939) | 6.6 (66,662) | 11.2 (79,397) | 9.6 (32,809) | 4.9 (18,071) |
| **Tuberculosis (ongoing episode)** | | | | | |
| Absent throughout 2020-2022 | 96.8 (2,351,220) | 96.7 (976,653) | 96.0 (682,764) | 96.8 (331,198) | 98.3 (360,605) |
| Experienced an episode during 2020-2022 | 3.2 (78,707) | 3.3 (33,019) | 4.0 (28,313) | 3.2 (11,110) | 1.7 (6,265) |
| **Pregnancy (ongoing episode)** | | | | | |
| Absent throughout 2020-2022 | 91.5 (2,222,462) | 82.4 (832,093) | 95.8 (681,306) | 100.0 (342,193) | 100.0 (366,870) |
| Experienced during 2020-2022 | 8.5 (207,465) | 17.6 (177,579) | 4.2 (29,771) | 0.0 (115) | 0.0 (0) |

| Characteristic | Overall<br>N = 2,429,927 | Age category (years) at start 2020<br>18-34<br>N = 1,009,672 | 35-49<br>N = 711,077 | 50-59<br>N = 342,308 | 60-99<br>N = 366,870 |
|---|---|---|---|---|---|
| **Number of negative SARS-CoV-2 tests (by end 2022)** | | | | | |
| 0 | 81.8 (1,987,267) | 83.1 (839,227) | 81.5 (579,659) | 79.8 (273,010) | 80.5 (295,371) |
| 1 | 13.1 (317,758) | 12.4 (124,958) | 13.2 (93,617) | 14.2 (48,662) | 13.8 (50,521) |
| 2+ | 5.1 (124,902) | 4.5 (45,487) | 5.3 (37,801) | 6.0 (20,636) | 5.7 (20,978) |
| **COVID-19 Vaccination status[b] and boosters (at end 2022)** | | | | | |
| None | 55.0 (1,336,826) | 60.9 (614,851) | 51.2 (363,908) | 49.0 (167,646) | 51.9 (190,421) |
| Incomplete BNT162b2 | 4.9 (118,559) | 6.2 (62,962) | 4.4 (31,589) | 3.6 (12,315) | 3.2 (11,693) |
| Complete Ad26.COV2S (no booster) | 9.7 (236,725) | 11.2 (112,933) | 12.3 (87,290) | 7.7 (26,221) | 2.8 (10,281) |
| Complete Ad26.COV2S (and booster) | 1.7 (41,196) | 1.4 (14,200) | 2.6 (18,363) | 1.9 (6,450) | 0.6 (2,183) |
| Complete BNT162b2 (no booster) | 22.6 (548,914) | 18.1 (183,198) | 24.3 (172,761) | 28.3 (96,908) | 26.2 (96,047) |
| Complete BNT162b2 (and booster) | 6.1 (147,707) | 2.1 (21,528) | 5.2 (37,166) | 9.6 (32,768) | 15.3 (56,245) |
| **Vital status (at end 2022)** | | | | | |
| Alive | 95.4 (2,318,859) | 98.7 (996,771) | 97.2 (690,943) | 94.0 (321,632) | 84.4 (309,513) |
| Died: not COVID-19 related | 4.1 (99,525) | 1.2 (12,399) | 2.6 (18,501) | 5.3 (18,277) | 13.7 (50,348) |
| Died: COVID-19 related | 0.5 (11,543) | 0.0 (502) | 0.2 (1,633) | 0.7 (2,399) | 1.9 (7,009) |
| **SARS-CoV-2 diagnosis (by end 2022)** | | | | | |
| Not experienced | 92.1 (2,237,396) | 93.4 (943,413) | 91.9 (653,256) | 90.1 (308,580) | 90.5 (332,147) |
| Experienced (2020-2022) | 7.9 (192,531) | 6.6 (66,259) | 8.1 (57,821) | 9.9 (33,728) | 9.5 (34,723) |
| **COVID-19 related hospitalisation or death (by end 2022)** | | | | | |
| Not experienced | 98.4 (2,391,586) | 99.4 (1,003,799) | 98.9 (702,936) | 97.6 (334,057) | 95.6 (350,794) |
| Experienced (2020-2022) | 1.6 (38,341) | 0.6 (5,873) | 1.1 (8,141) | 2.4 (8,251) | 4.4 (16,076) |
| **COVID-19 related hospitalisation with severe disease or death (by end 2022)** | | | | | |
| Not experienced | 99.3 (2,412,279) | 99.9 (1,008,359) | 99.6 (707,897) | 98.8 (338,362) | 97.5 (357,661) |
| Experienced (2020-2022) | 0.7 (17,648) | 0.1 (1,313) | 0.4 (3,180) | 1.2 (3,946) | 2.5 (9,209) |
| **COVID-19 related death (by end 2022)** | | | | | |
| Not experienced | 99.5 (2,418,384) | 100.0 (1,009,170) | 99.8 (709,444) | 99.3 (339,909) | 98.1 (359,861) |
| Experienced (2020-2022) | 0.5 (11,543) | 0.0 (502) | 0.2 (1,633) | 0.7 (2,399) | 1.9 (7,009) |

PHC: Primary healthcare

[a] Tuberculosis (at any time in past) includes those with an ongoing episode.

[b] Complete vaccination: ≥28 days since a first Ad26.COV2.S dose or ≥14 days since a second dose following a first BNT162b2 dose; incomplete BNT162b2: ≥21 days since a first BNT162b2 dose but not yet completely vaccinated (see methods).

### B2. COVID-19 outcomes, SARS-CoV-2 testing and positivity, and COVID-19 vaccination over time

The figures reported in this section are similar to Figure 1 of the article. However, here we consider distinct (sub)cohorts within our study population, as defined by sex, age at the start of 2020, and HIV status at the start of 2020.

Note that the scales of the y-axes may vary by stratification variable.

Figure footnotes:

[a] Proportions are not shown for first and last months due to large uncertainties from few diagnoses/tests.

[b] A person is completely vaccinated ≥28 days since a first Ad26.COV2S dose or ≥14 days since a second dose following a first BNT162b2 dose; and is in the incomplete BNT162b2 state ≥21 days since a first BNT162b2 dose and while not yet completely vaccinated. 'No vaccine / transition' means the person has not received a dose or it is <28 days since a first Ad26.COV2S dose or <21 days since a first BNT162b2 dose. The time in the current vaccinated state (i.e. time since entering the state, which is reset 7 days following a booster dose) is also shown (< 6 versus ≥ 6 months). See methods of the article.

#### *B2.1 Outcomes, testing and vaccination over time: Males*

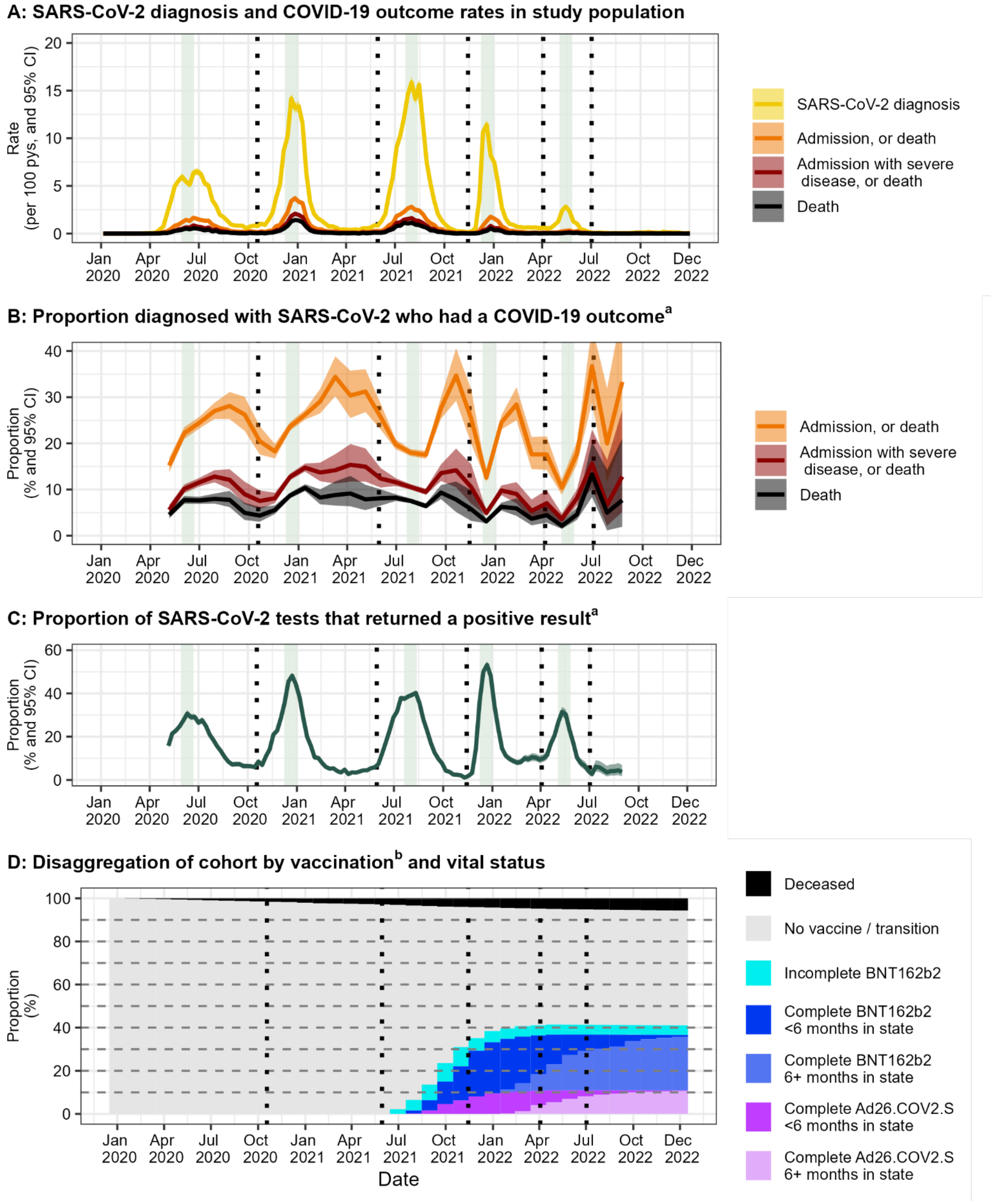

#### *B2.2 Outcomes, testing and vaccination over time: Females*

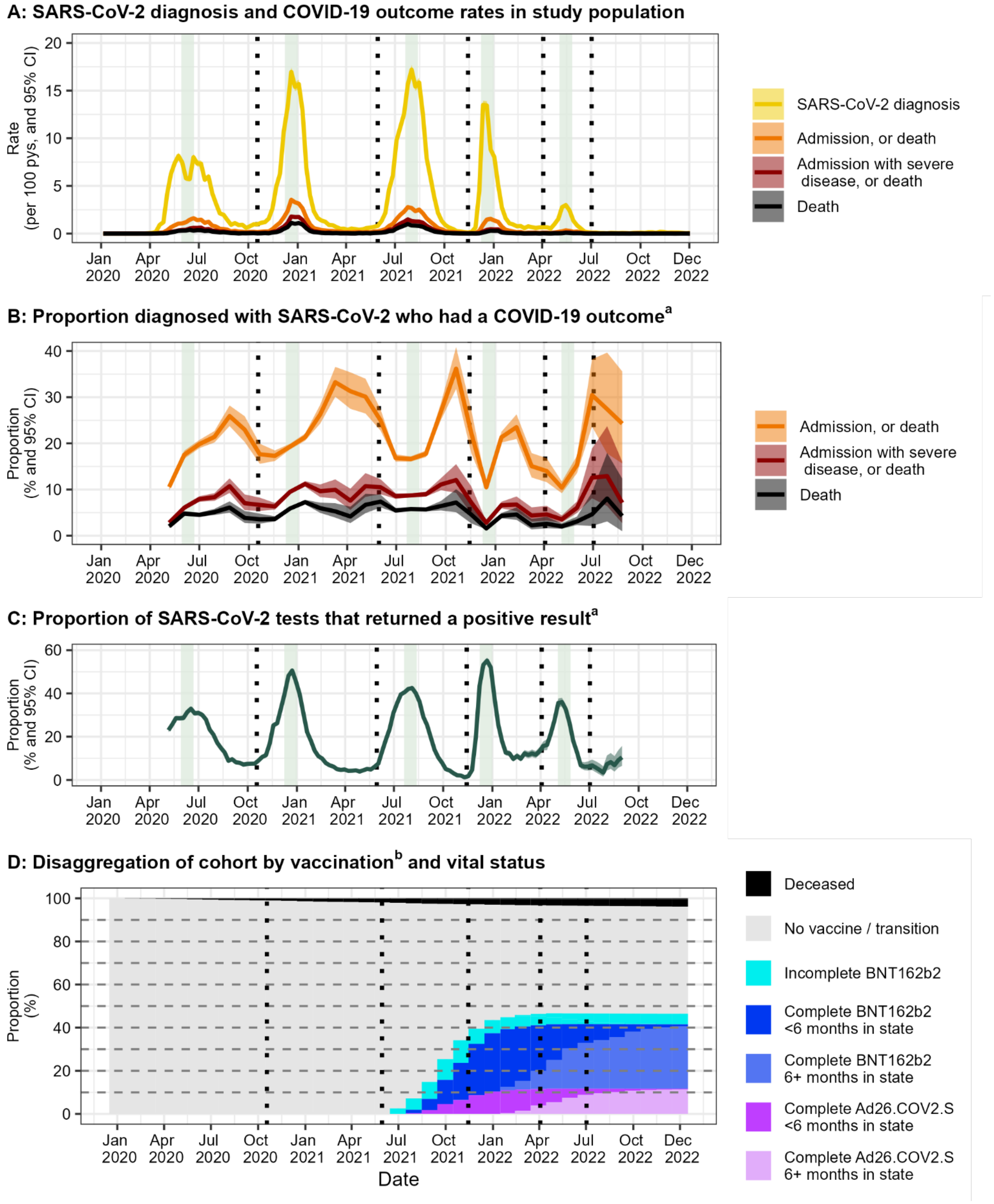

#### *B2.3 Outcomes, testing and vaccination over time: Age 18-34 years*

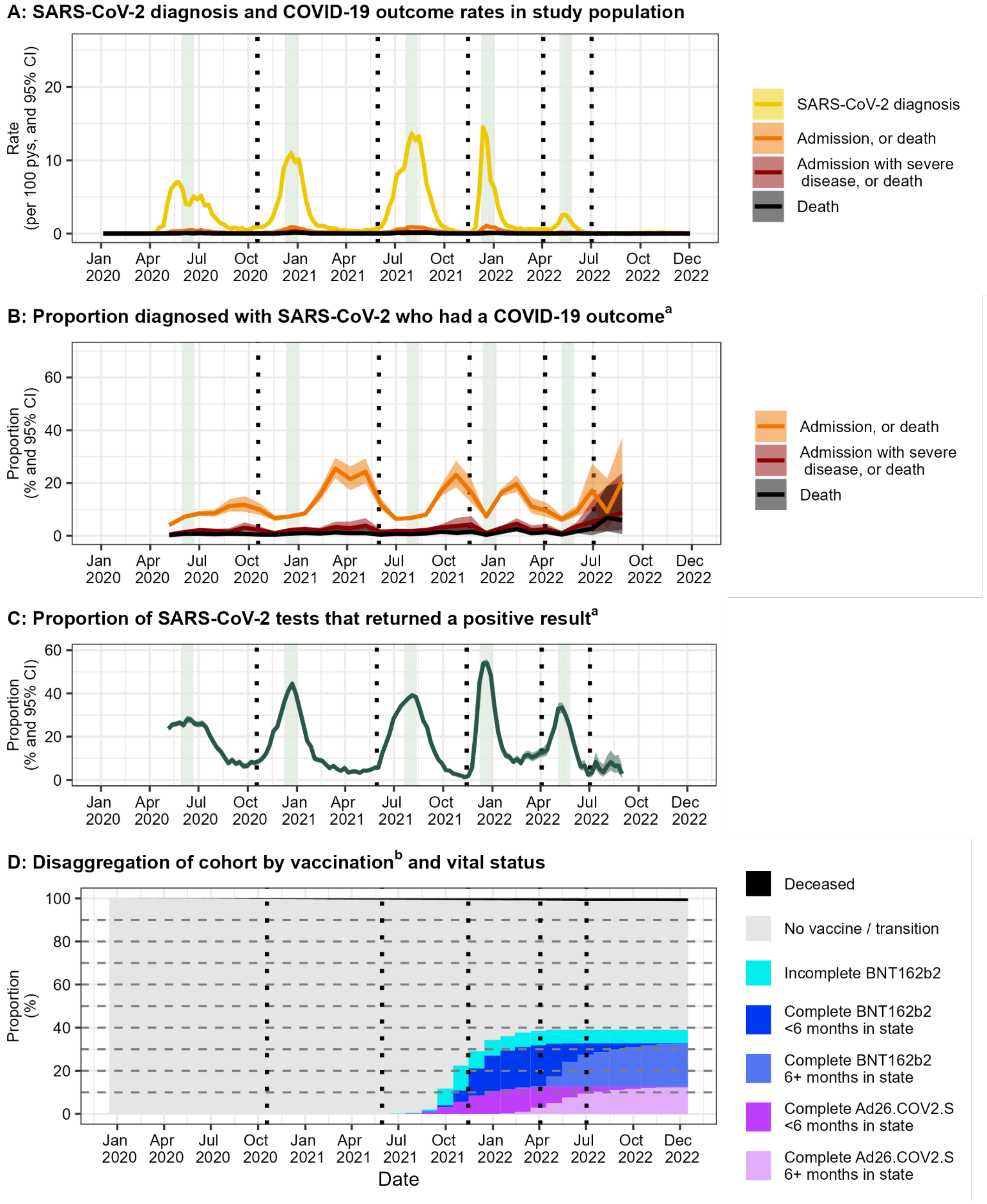

#### *B2.4 Outcomes, testing and vaccination over time: Age 35-49 years*

**A: SARS-CoV-2 diagnosis and COVID-19 outcome rates in study population**

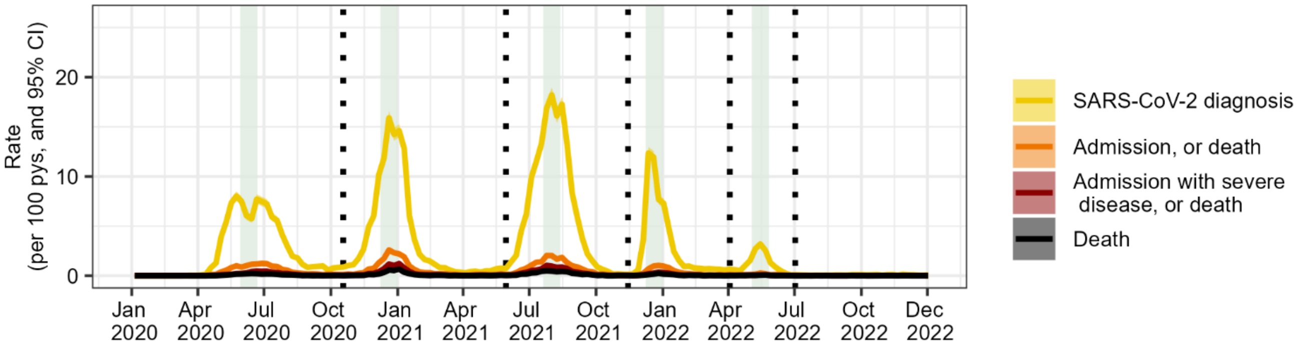


**B: Proportion diagnosed with SARS-CoV-2 who had a COVID-19 outcome[a]**

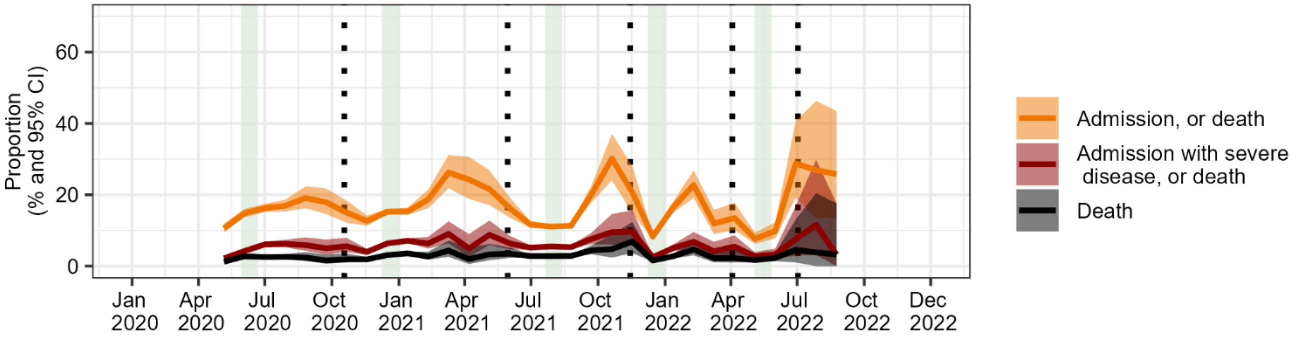


**C: Proportion of SARS-CoV-2 tests that returned a positive result[a]**

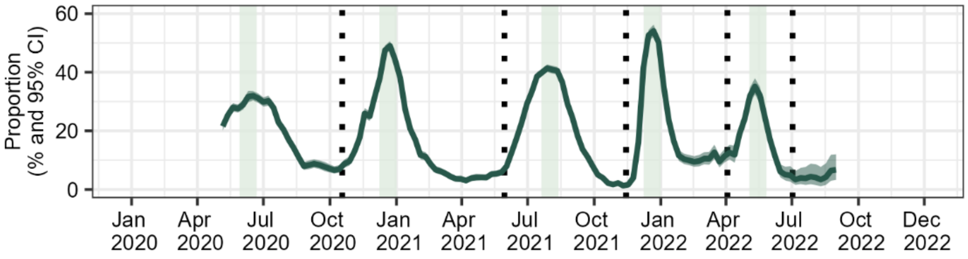


**D: Disaggregation of cohort by vaccination[b] and vital status**

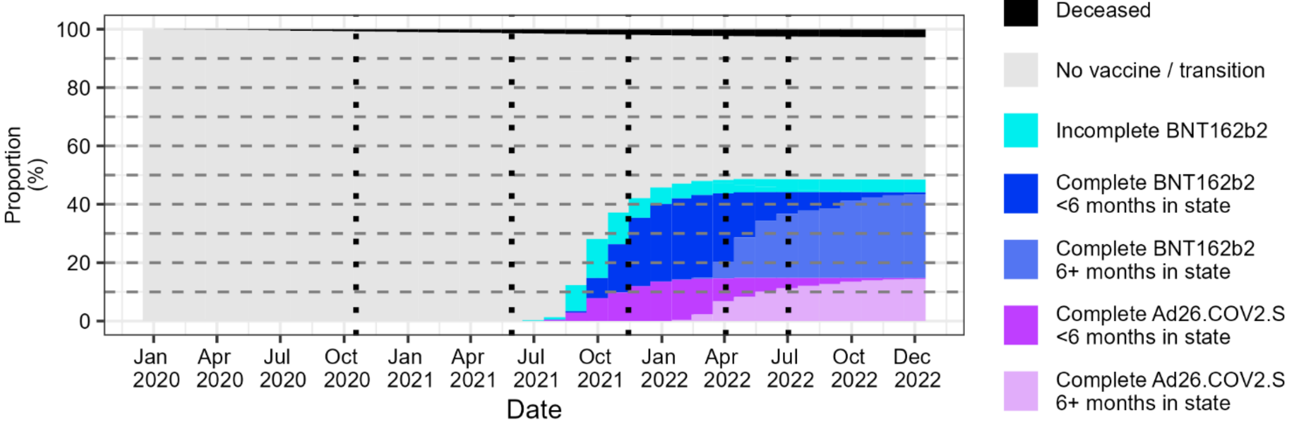

#### *B2.5 Outcomes, testing and vaccination over time: Age 50-59 years*

**A: SARS-CoV-2 diagnosis and COVID-19 outcome rates in study population**

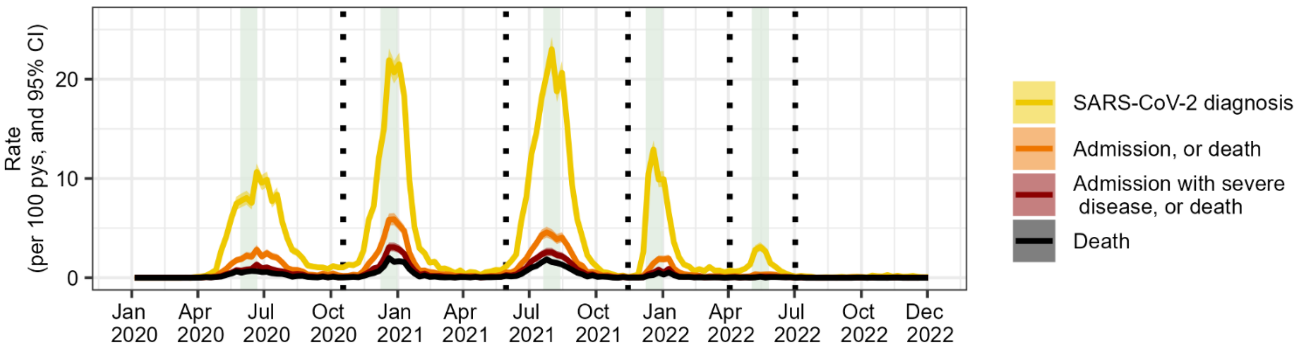


**B: Proportion diagnosed with SARS-CoV-2 who had a COVID-19 outcome[a]**

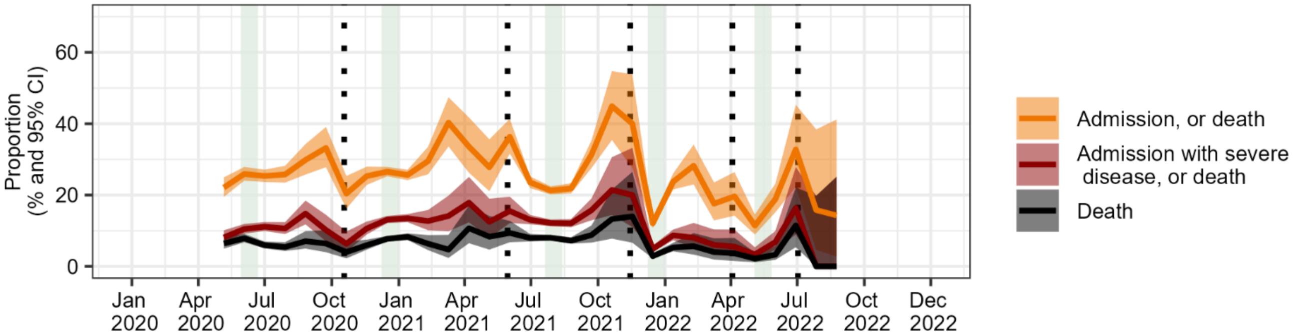


**C: Proportion of SARS-CoV-2 tests that returned a positive result[a]**

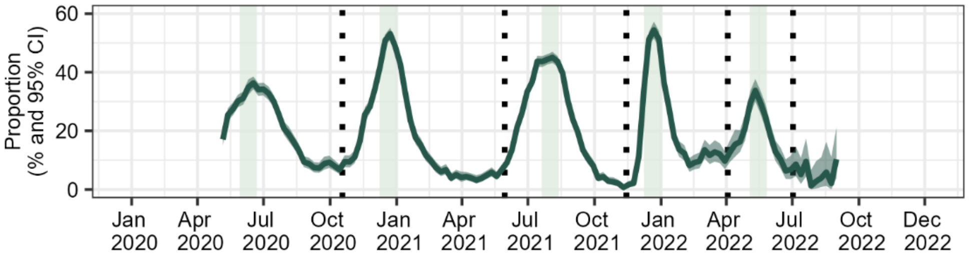


**D: Disaggregation of cohort by vaccination[b] and vital status**

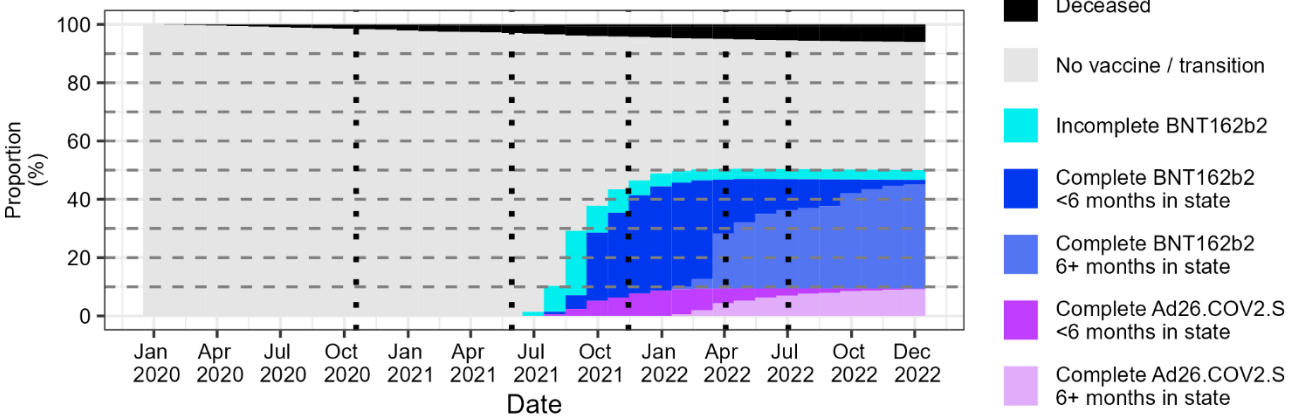

#### *B2.6 Outcomes, testing and vaccination over time: Age 60+ years*

**A: SARS-CoV-2 diagnosis and COVID-19 outcome rates in study population**

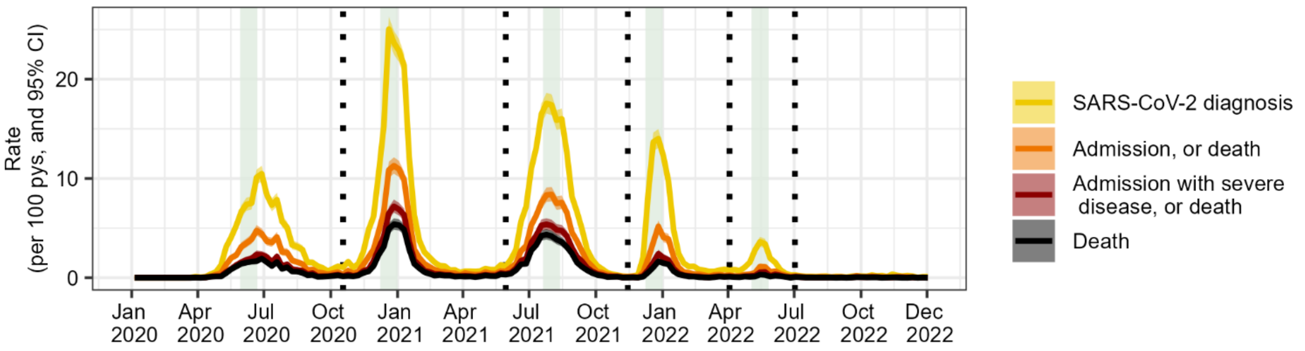


**B: Proportion diagnosed with SARS-CoV-2 who had a COVID-19 outcome[a]**

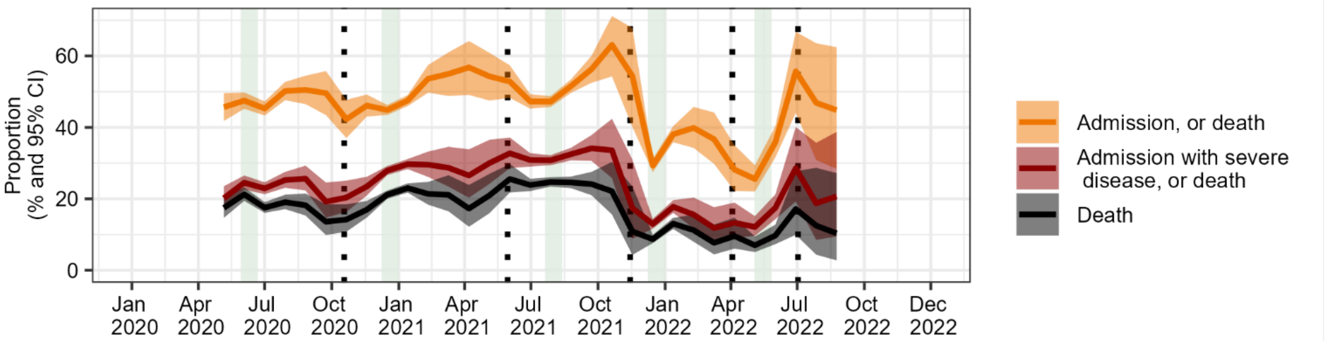


**C: Proportion of SARS-CoV-2 tests that returned a positive result[a]**

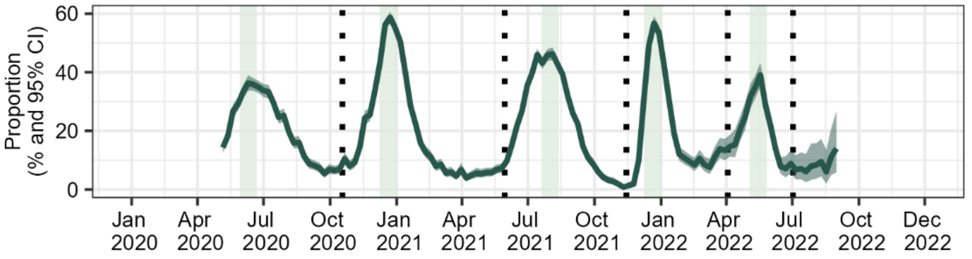


**D: Disaggregation of cohort by vaccination[b] and vital status**

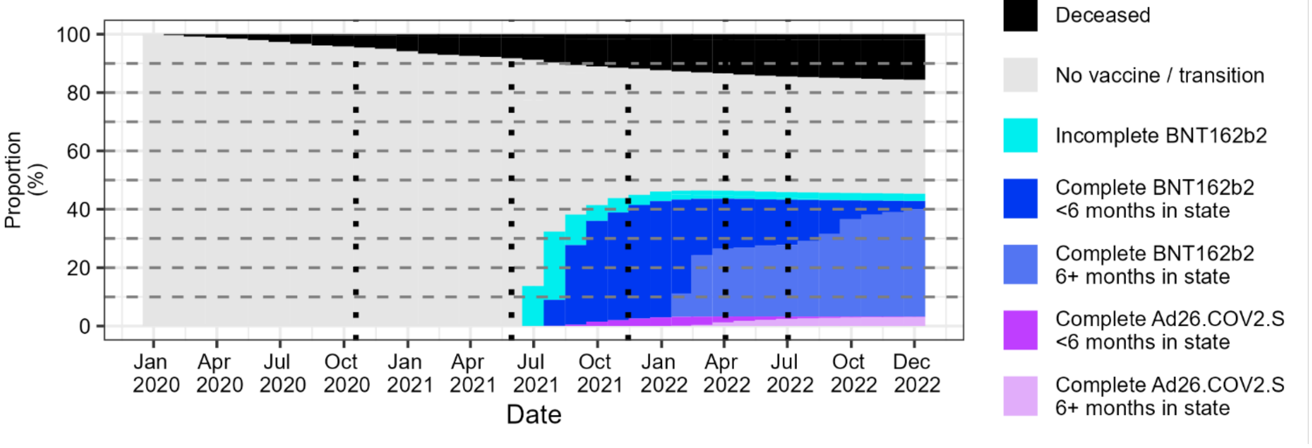

### *B2.7 Outcomes, testing and vaccination over time: Living with HIV at start 2020*

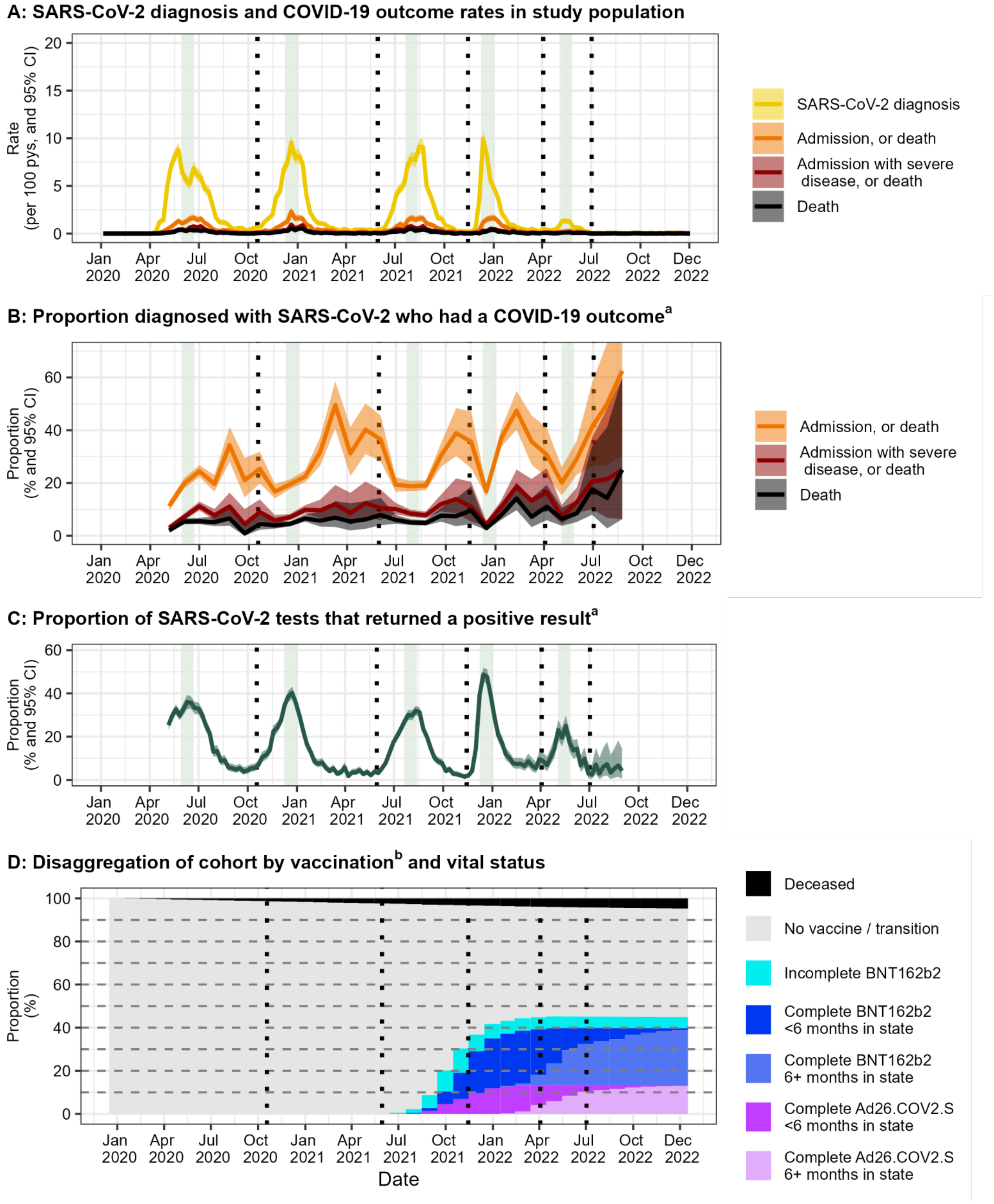

#### *B2.8 Outcomes, testing and vaccination over time: Not known to be living with HIV at start 2020*

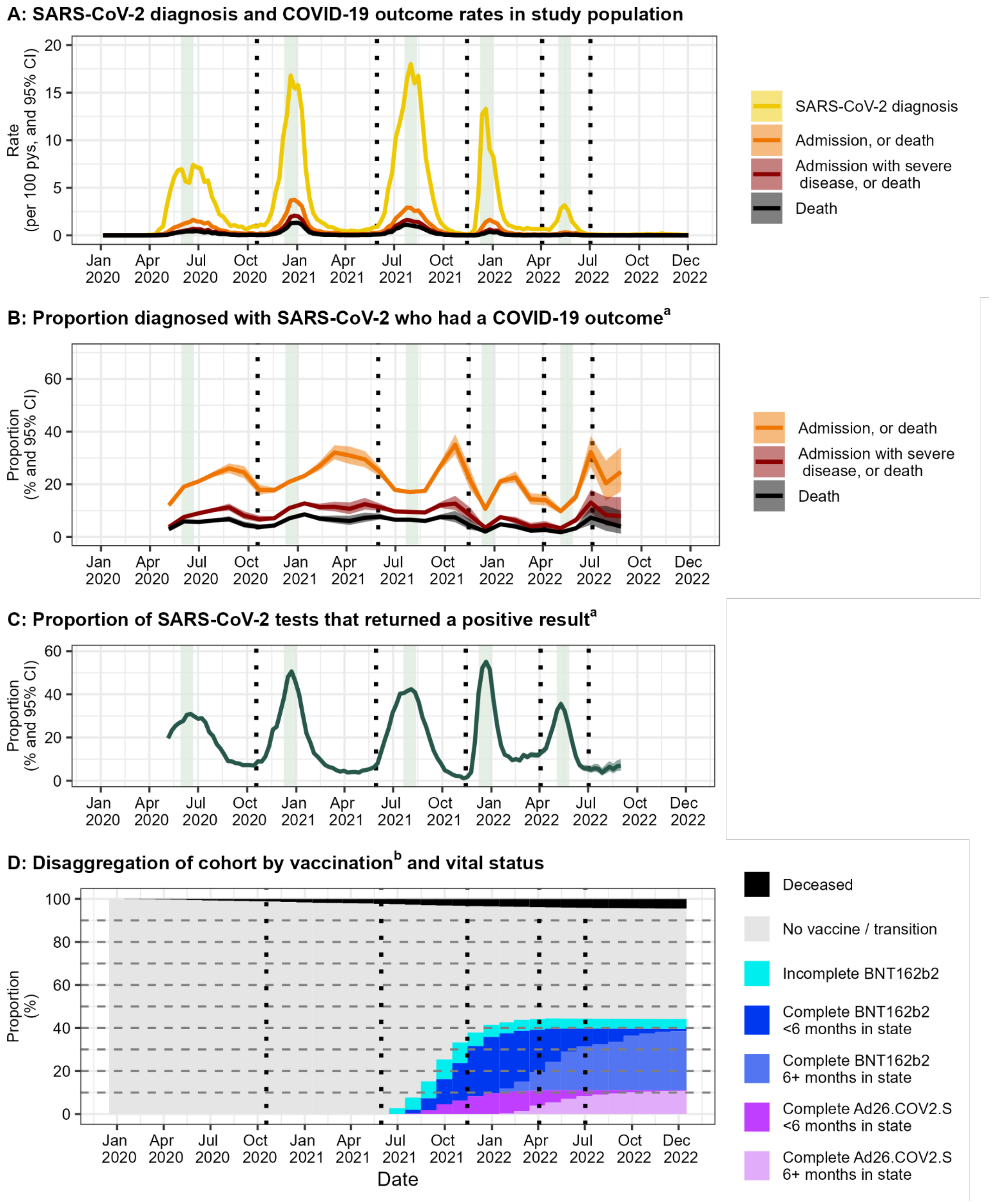




See Kassanjee et al. COVID-19 vaccine uptake and effectiveness by time since vaccination in the Western Cape province, South Africa: An observational cohort study during 2020-2022.

### B3. Vaccine effectiveness for distinct subpopulations

The figures reported in this section are similar to Figure 3 in the article.

However, here we consider distinct (sub)cohorts within our study population, as defined by sex, age at the start of 2020 and HIV status at the start of 2020. We also report vaccine effectiveness when excluding people who received heterologous doses, from the time they received their first dose.

#### *B3.1 Vaccine effectiveness: Males*

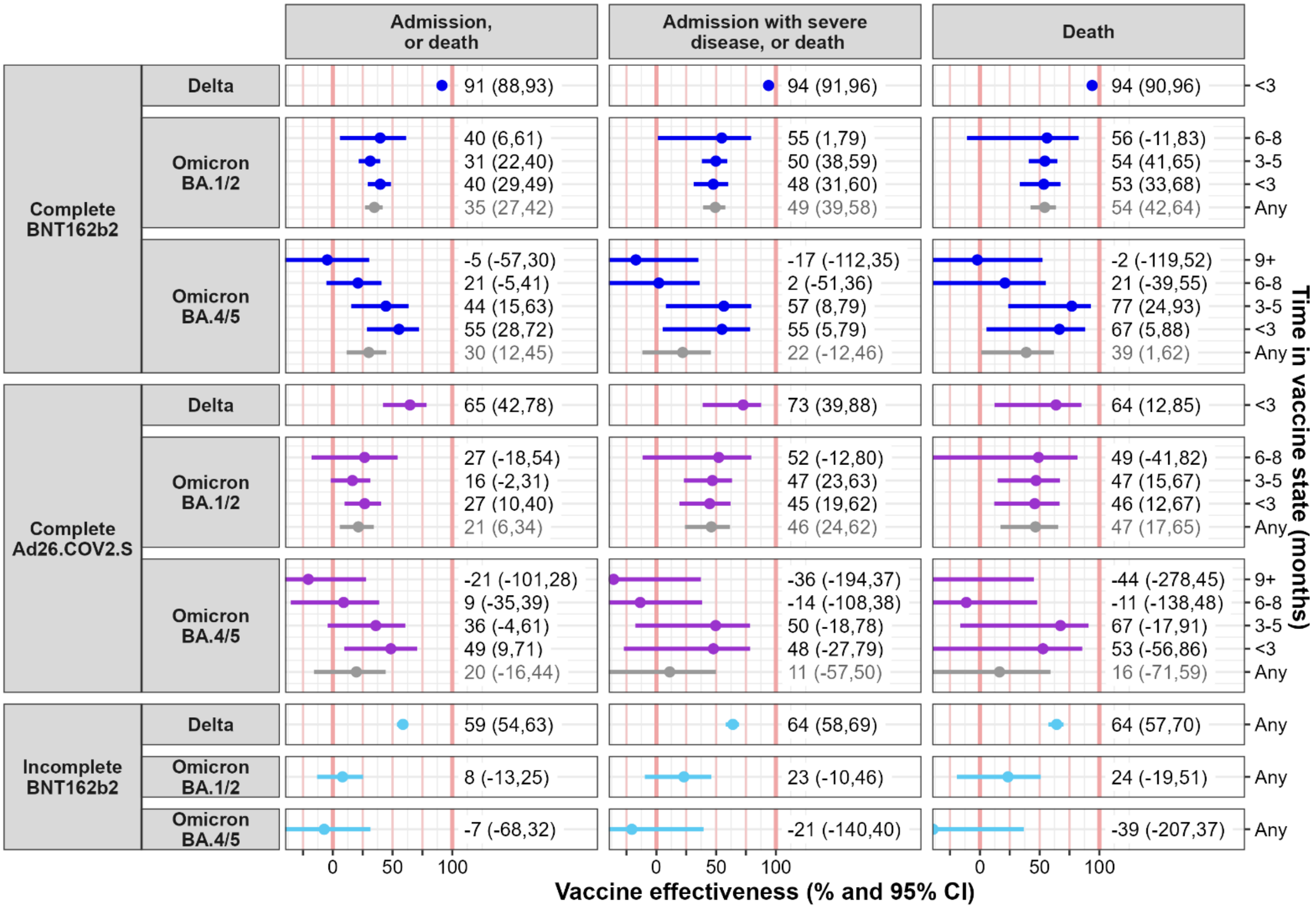

#### *B3.2 Vaccine effectiveness: Females*

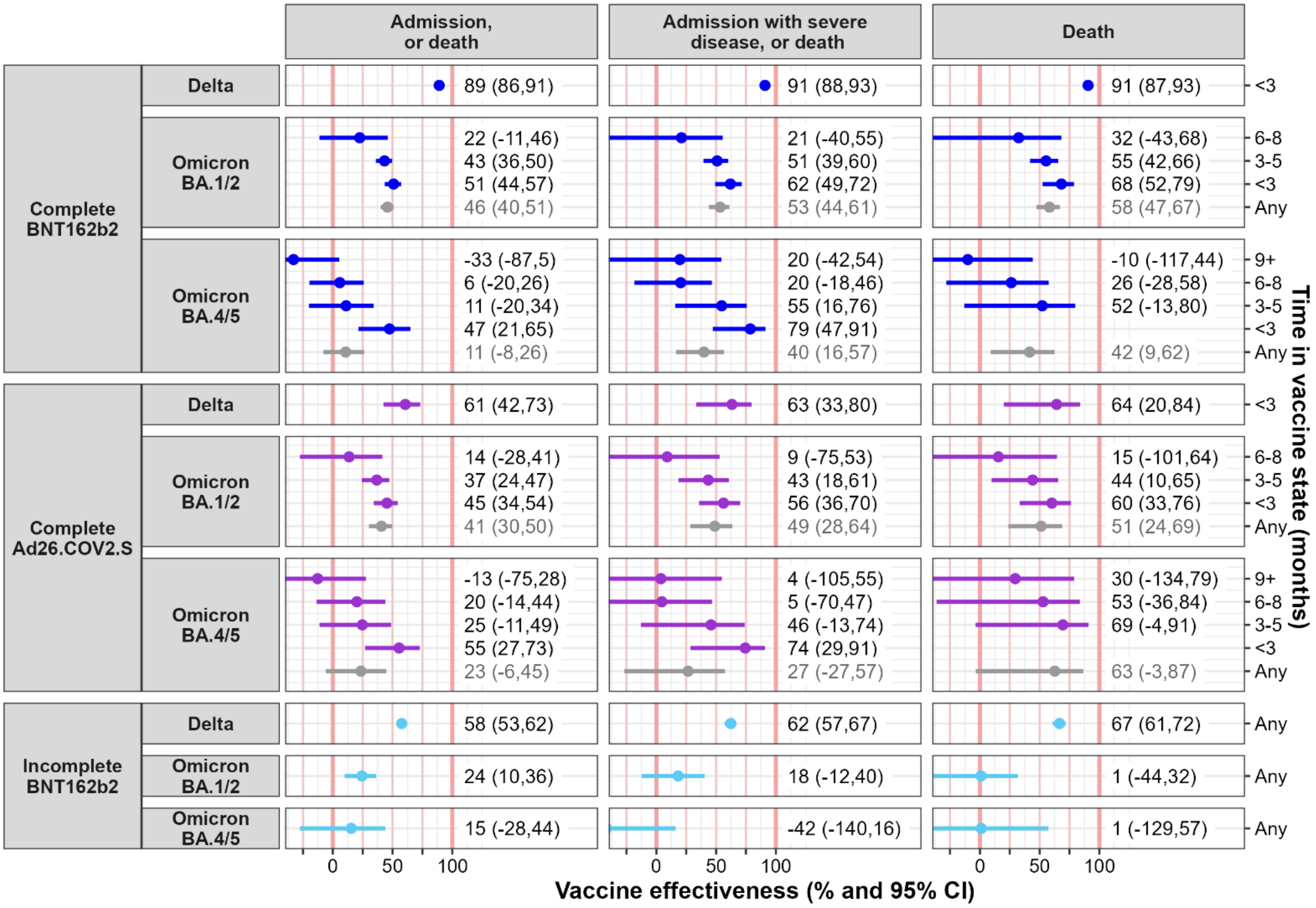

#### *B3.3 Vaccine effectiveness: Age 18-49 years*

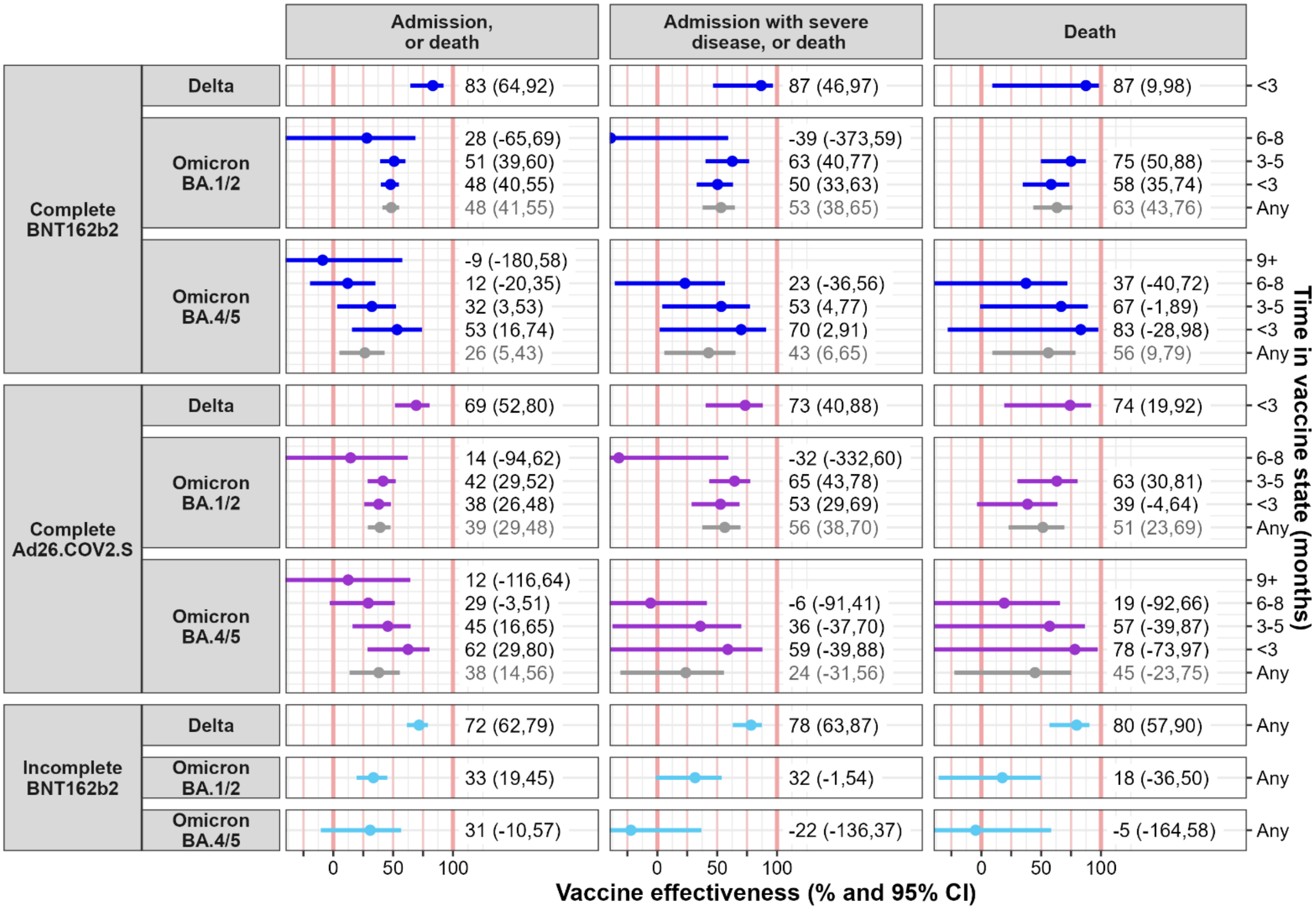

#### *B3.4 Vaccine effectiveness: Age 50+ years*

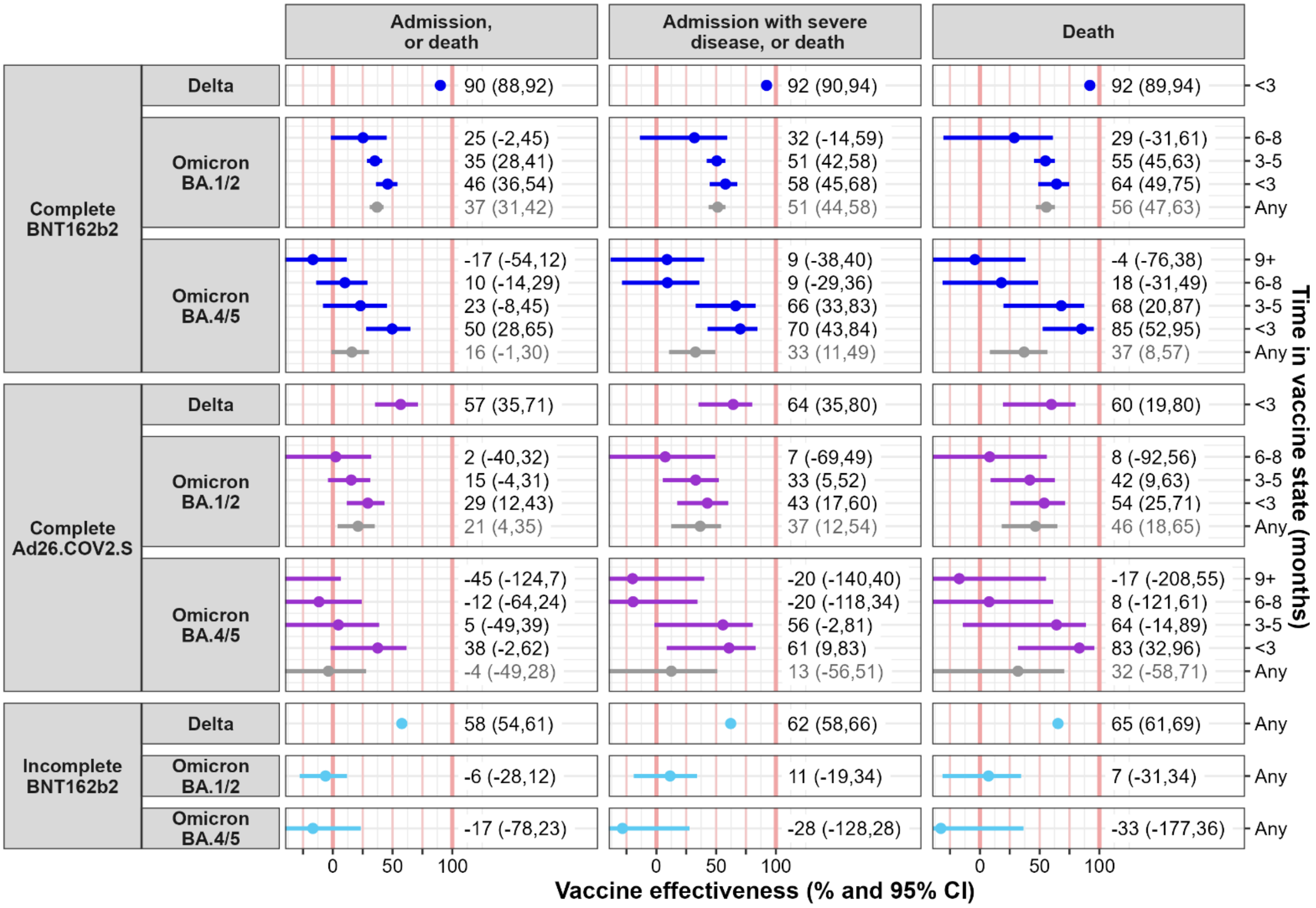

#### *B3.5 Vaccine effectiveness: Known to be living with HIV at start 2020*

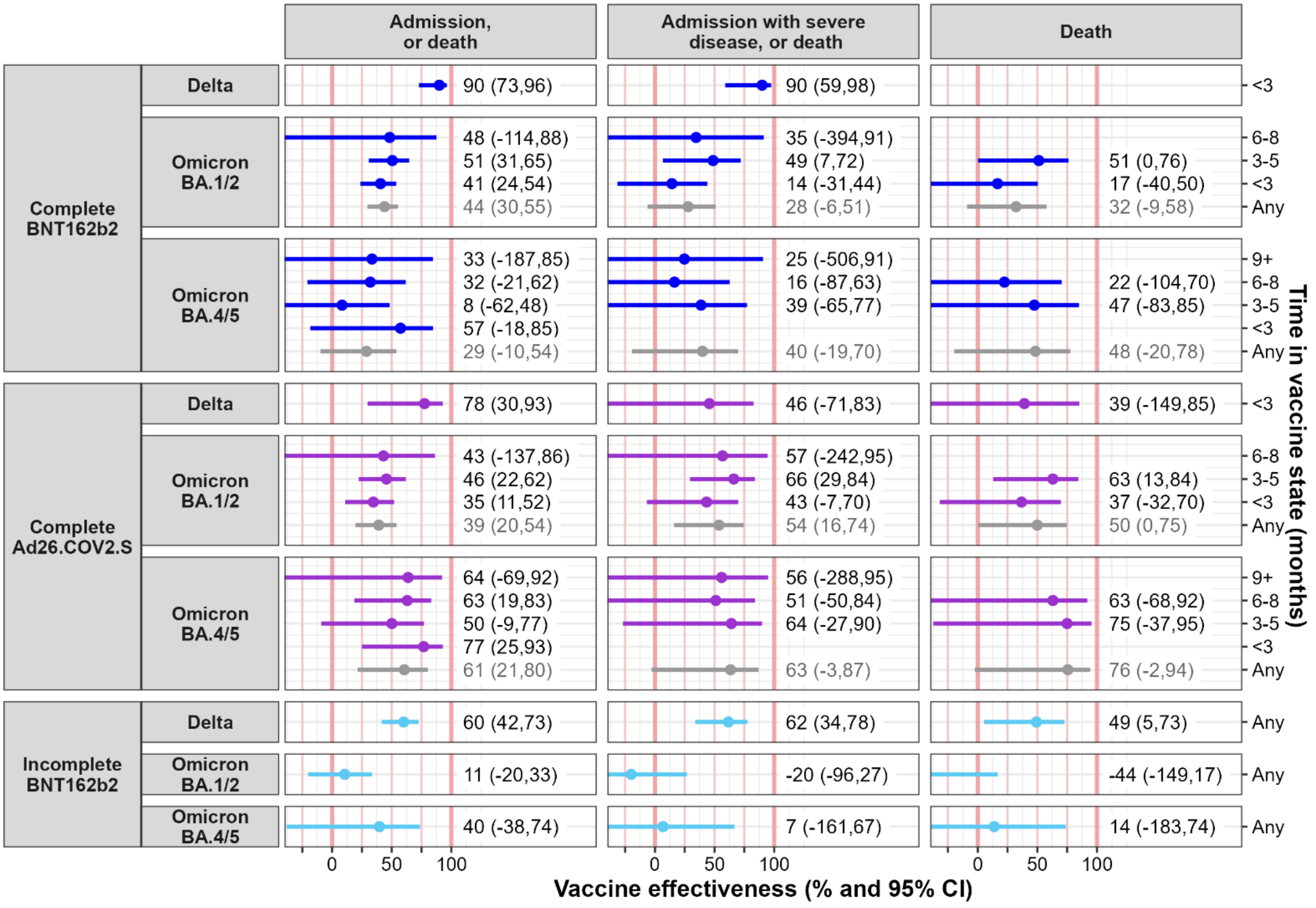

#### *B3.6 Vaccine effectiveness: Not known to be living with HIV at start 2020*

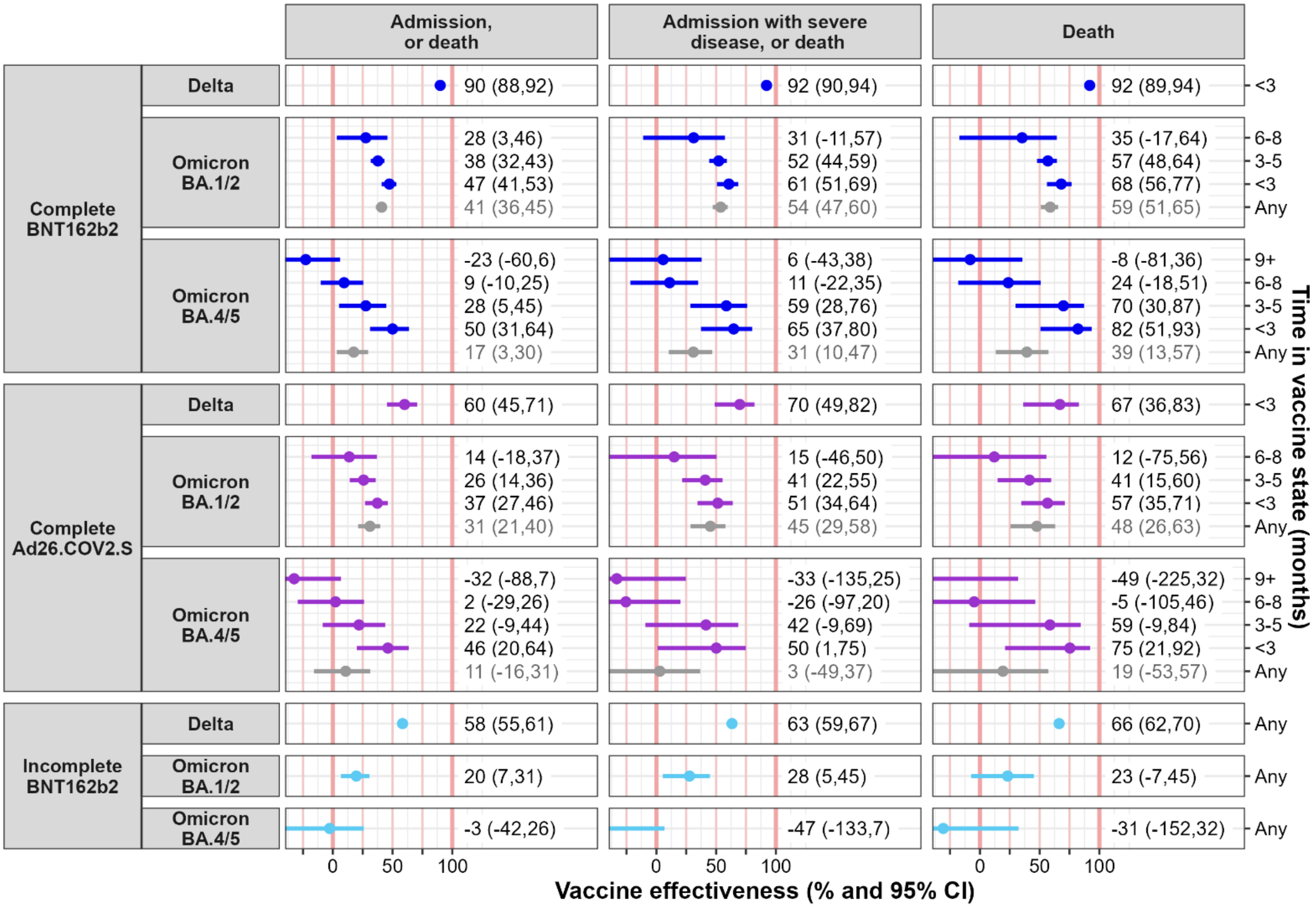

#### *B3.7 Vaccine effectiveness: Excluding people who received heterologous vaccines*

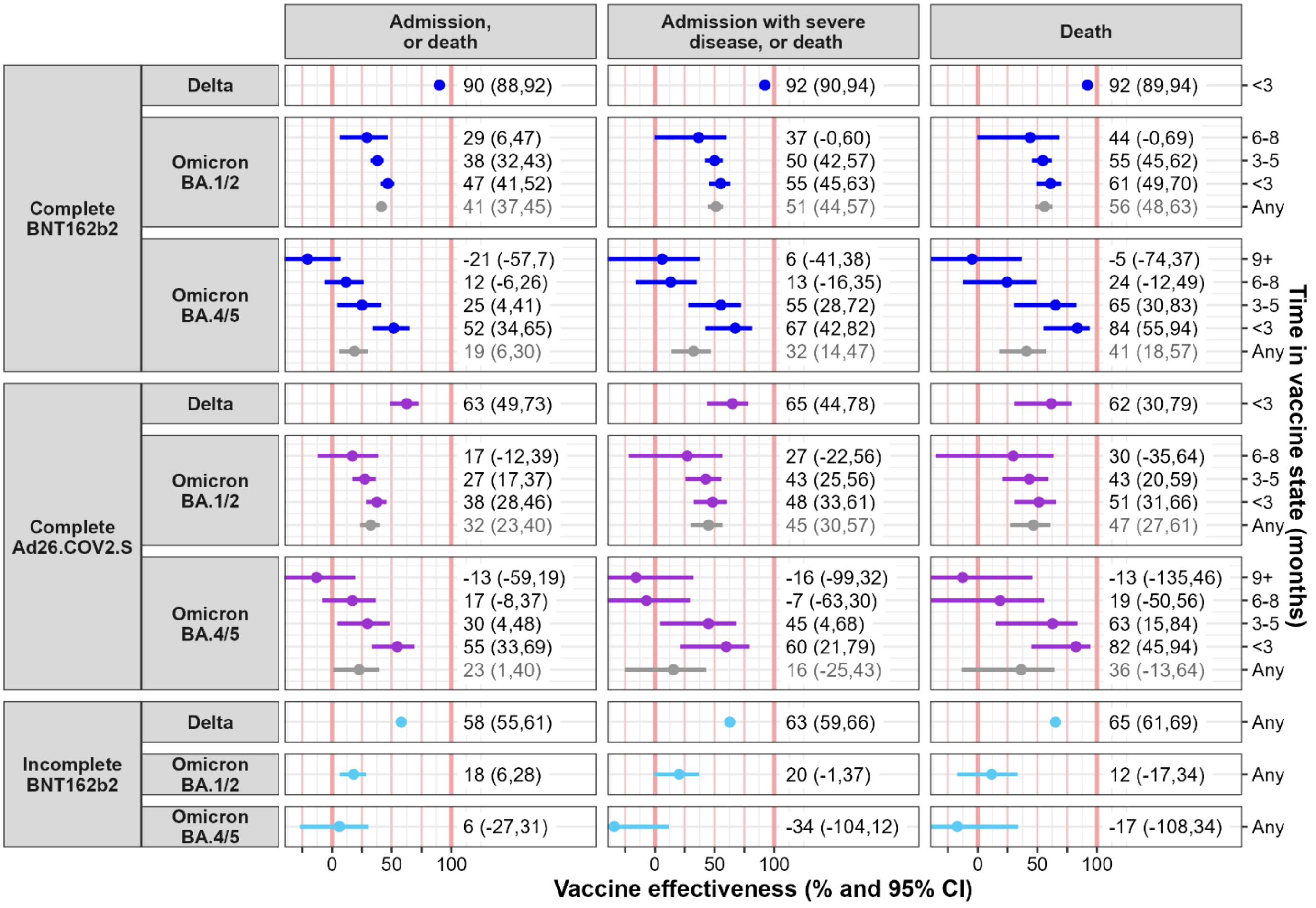


People who received heterologous doses were excluded from the time of receiving their first dose.

Among those people alive at start 2020 who completed their BNT162b2 primary vaccination series by end 2022, 28 of 696,621 (0.004%) persons received a heterologous second dose. Considering all 1,093,101 people who received at least one dose by end 2022 (incomplete or complete primary vaccination series, with or without boosters), 1.43% received heterologous doses (28 people as part of the primary series, and the rest during boosting).

### B4. All multivariable cox model hazard ratios

For completeness of reporting, the fitted hazard ratios for all model terms (vaccination terms and covariates) in the multivariable cox model used to estimate vaccine effectiveness are reported below. All hazard ratios reported in the section are from the same multivariable model, which adjust for all covariates.

For each set of model terms, the hazard ratio (x-axis) is shown for the different model terms (figure rows) and COVID-19 outcomes (figure columns), for the three wave periods that occurred after vaccine rollout (legend). The hazard ratio is the rate of the outcome in a category of persons, relative to a reference category (as indicated in the figure row label), holding all other model terms fixed. Hazard ratios greater than 1 represent higher outcome rates; and less than 1 lower outcome rates.

The focus of the article is on reporting the vaccine effectiveness estimates. The direct interpretation of the hazard ratios for the covariates presented below are complex and outside of the focus of this work: for example, a higher rate out of an outcome in a category of persons may results from more exposure to the virus (e.g. limited opportunity to socially distance), a greater probability of COVID-19 disease (or more severe disease) given that the person has acquired the SARS-CoV-2 virus, greater SARS-CoV-2 testing, or an association of COVID-19 outcomes with a variable for which the covariate is only a proxy or associated variable.

#### *B4.1 Hazard ratios: Demographic characteristics*

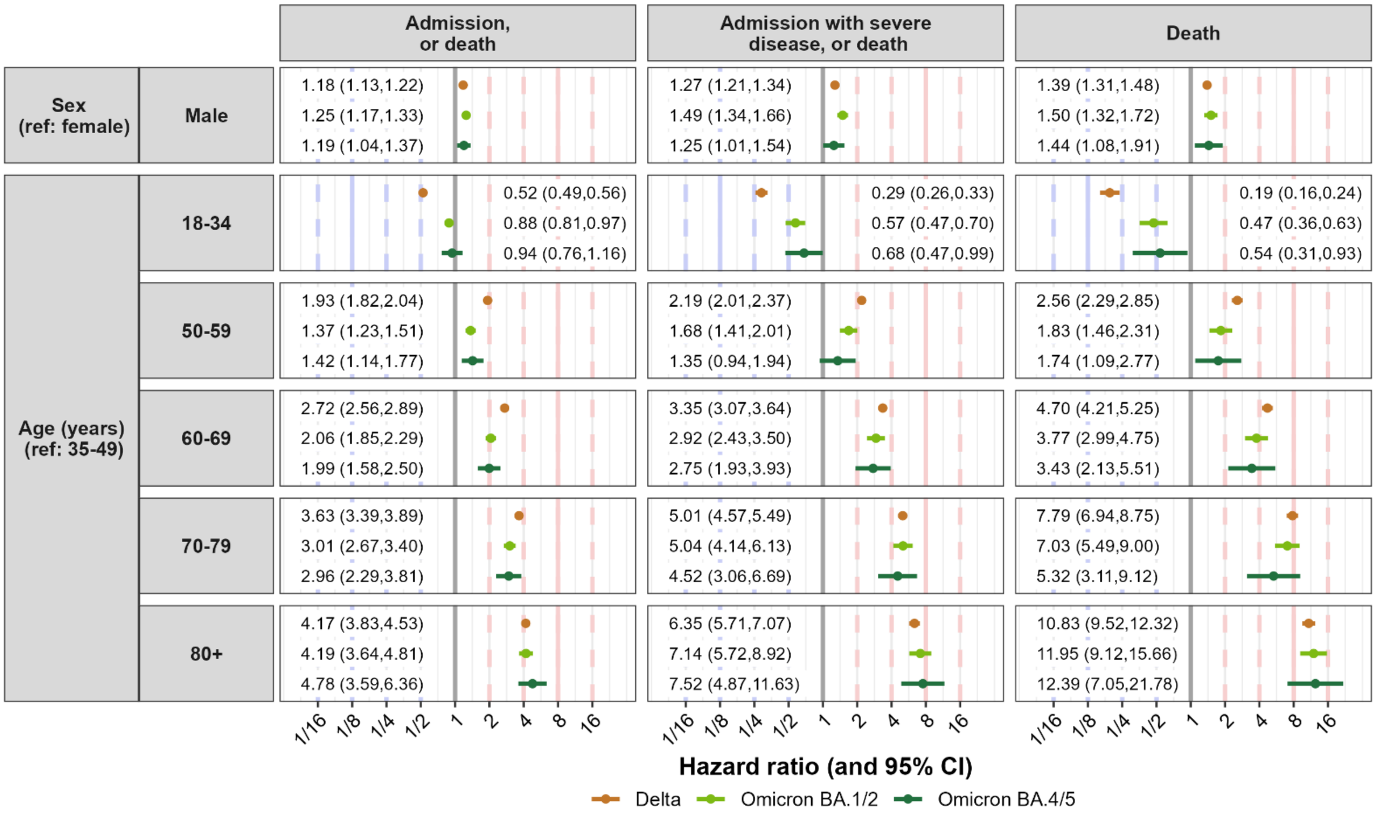

#### B4.2 Hazard ratios: Non-communicable conditions

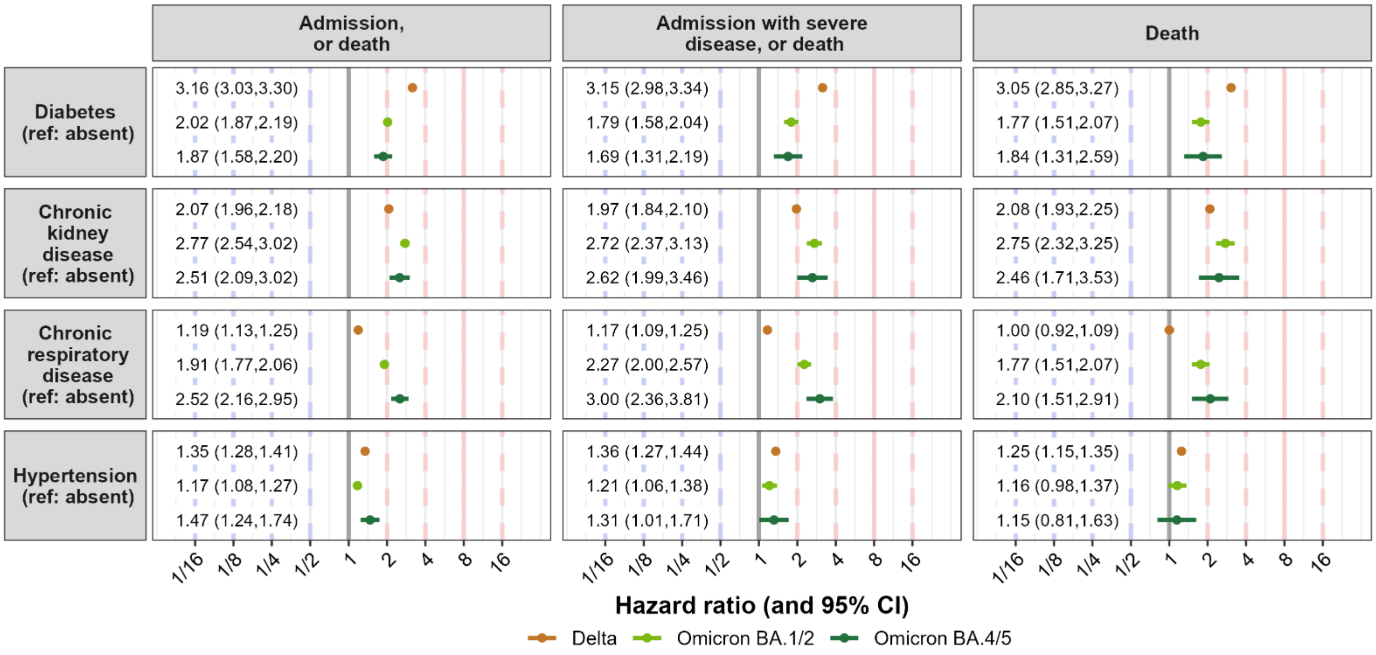


#### B4.3 Hazard ratios: Infectious conditions

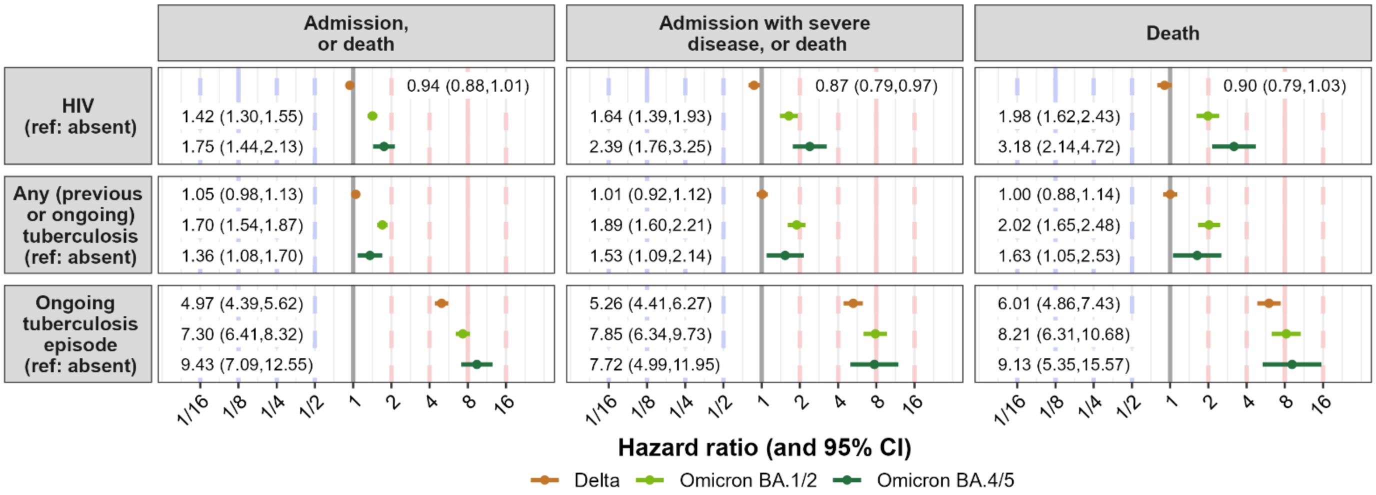


'Any' tuberculosis includes those with a previous or ongoing episode – i.e. the effect of an 'ongoing tuberculosis episode' is the *additional* effect of an episode being ongoing rather than only at some point in the past.

#### *B4.4 Hazard ratios: Other conditions*

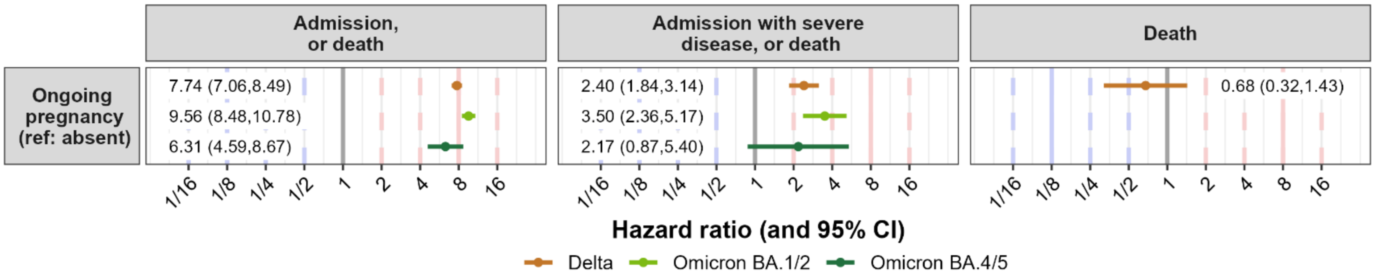


#### *B4.5 Hazard ratios: Prior SARS-CoV-2 diagnosis*

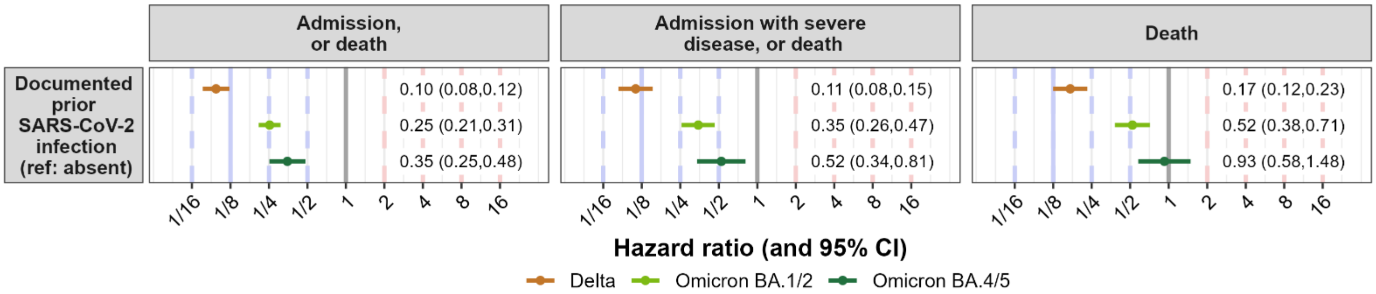

#### *B4.6 Hazard ratios: Location*

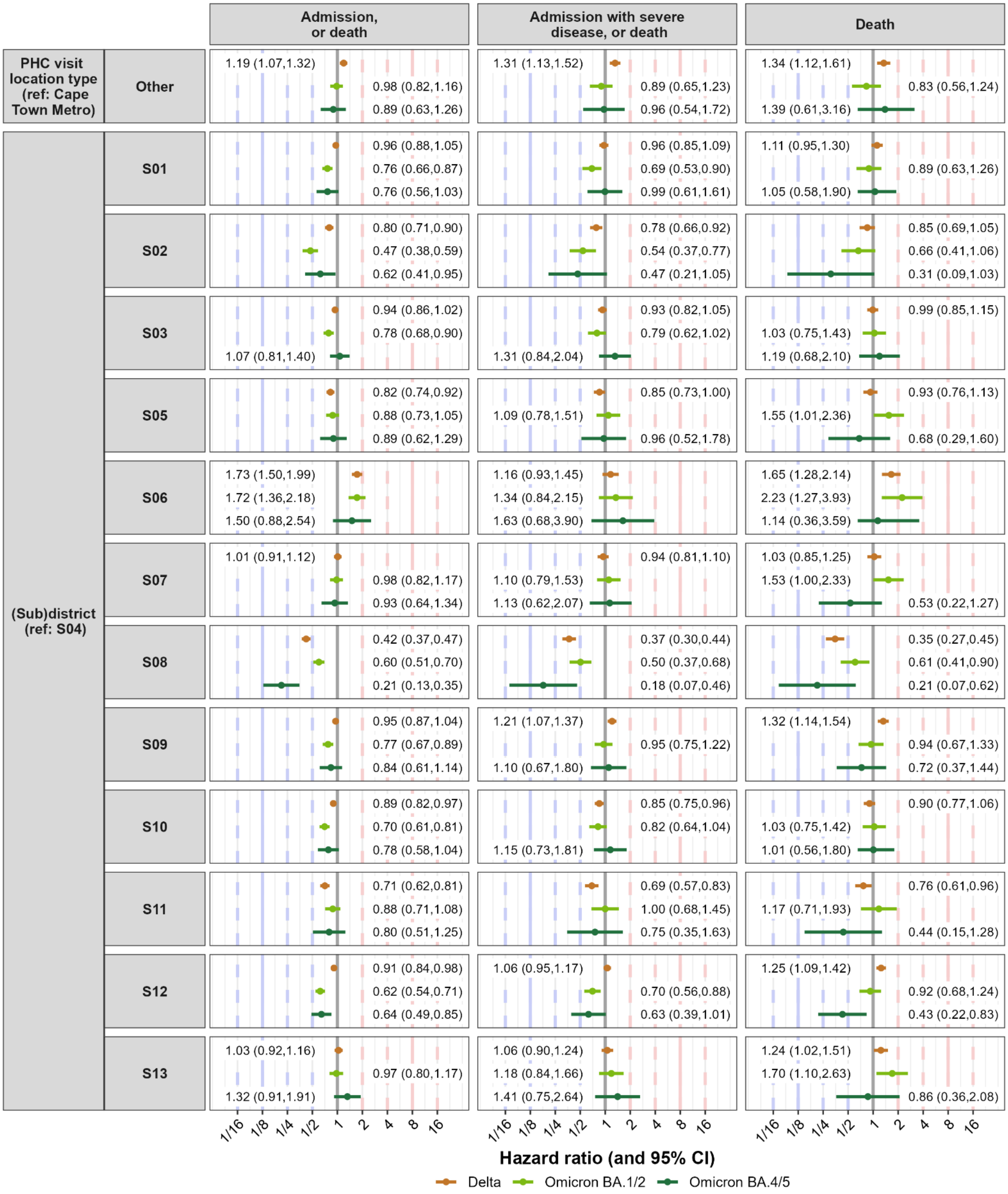


For PHC (primary healthcare) visit location type, 'Other' includes non-Metro and no recorded visit.

#### *B4.7 Hazard ratios: Healthcare utilisation and testing*

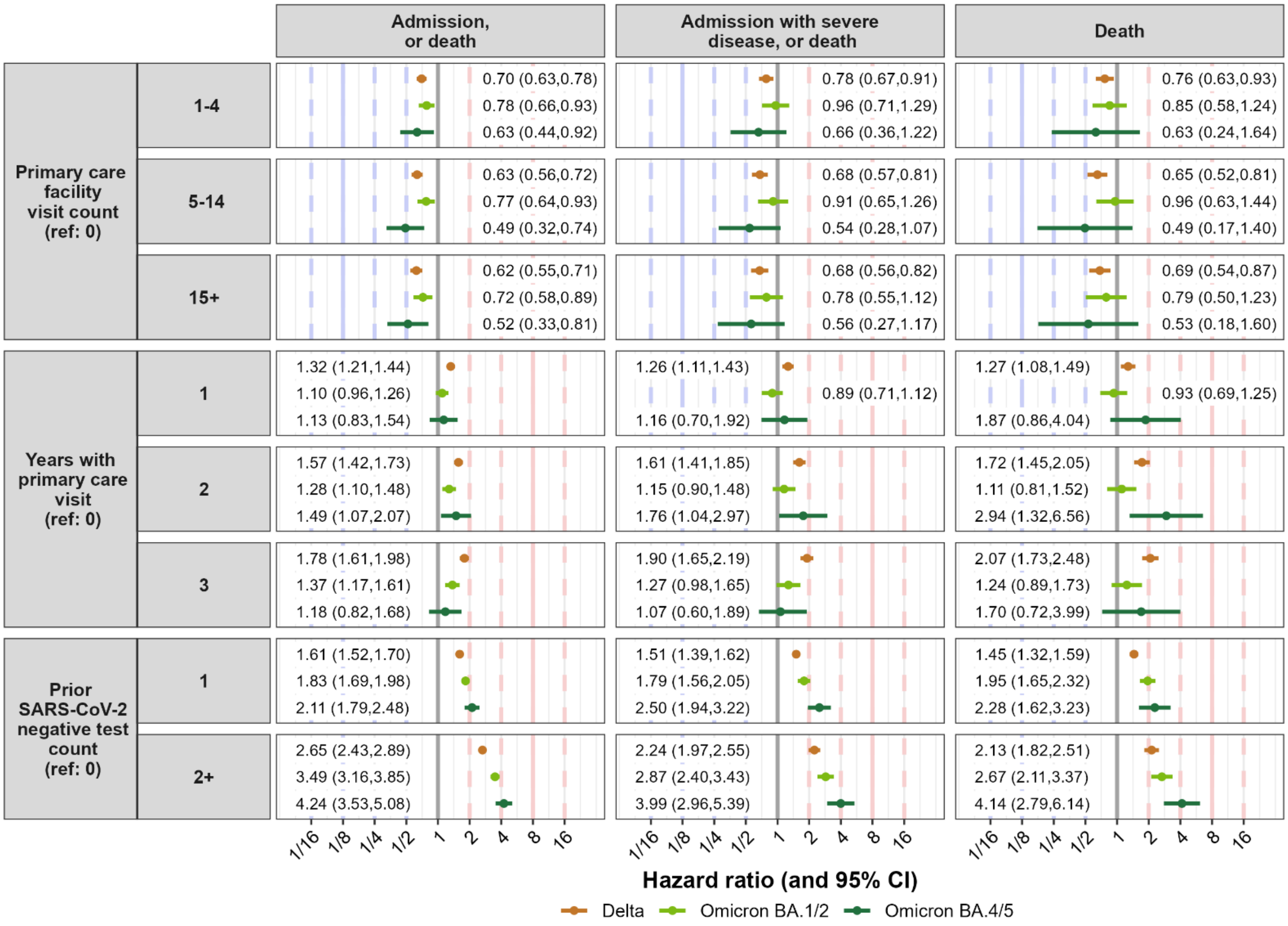

#### *B4.8 Hazard ratios: Vaccine terms*

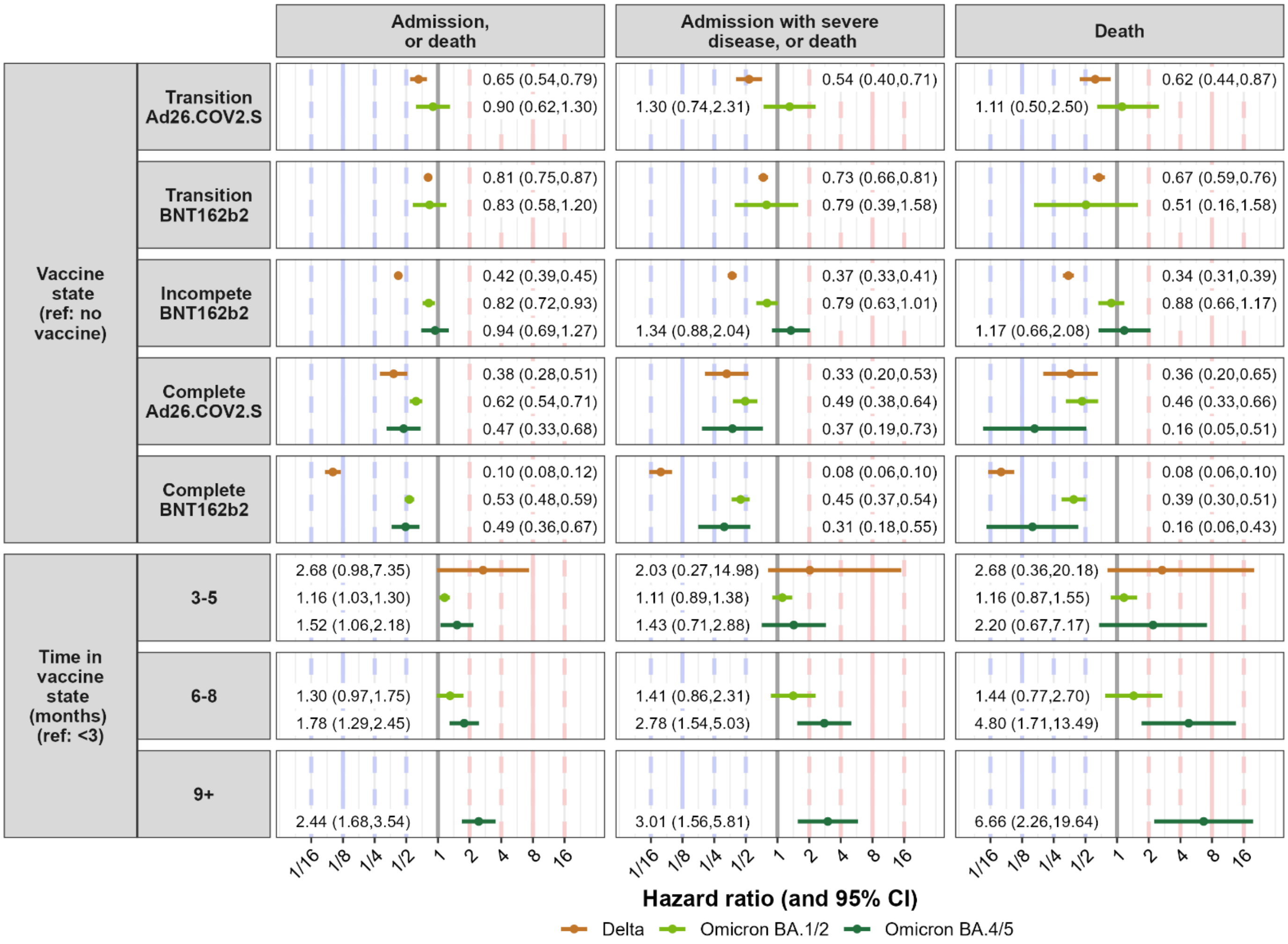

### B5. Vaccine effectiveness for rolling windows of calendar time

To understand how the associations between vaccine status and COVID-19 outcomes change over calendar time, the multivariable cox model was fitted to rolling windows of calendar time – each of duration 6 weeks, and each window starting 3 weeks after the previous window began. Due to limited precision when using data from short windows of time, vaccine state was not further distinguished by time in the vaccine state (time in vaccine state was included in the primary model reported in the article).

Changes in associations of vaccination with severe COVID-19 over calendar time are likely due to many factors, such as variations in time since most recent vaccine dose, testing coverage and match of vaccine and dominant infection (sub)lineage.

The figure below reports vaccine effectiveness estimates (x-axis) for each vaccinated state (figure rows) and COVID-19 outcome (figure columns), over time (y-axis, midpoint of window). The horizontal shaded areas (orange, light green, dark green) correspond to the dominant (sub)lineage in different periods (i.e., the distinct analysis wave periods used in the primary analysis). The grey curved line represents a locally weighted smoothing of the trend, to informally guide the eye. Vaccine effectiveness is 1 minus the ratio of rates of outcomes in vaccinated persons versus unvaccinated persons, expressed as a percentage: i.e. 100% is perfect 'protection' and 0% is no 'protection', shown by vertical red lines.

A person is completely vaccinated ≥28 days since a first Ad26.COV2S dose or ≥14 days since a second dose following a first BNT162b2 dose; and is in the incomplete BNT162b2 state ≥21 days since a first BNT162b2 dose and while not yet completely vaccinated.

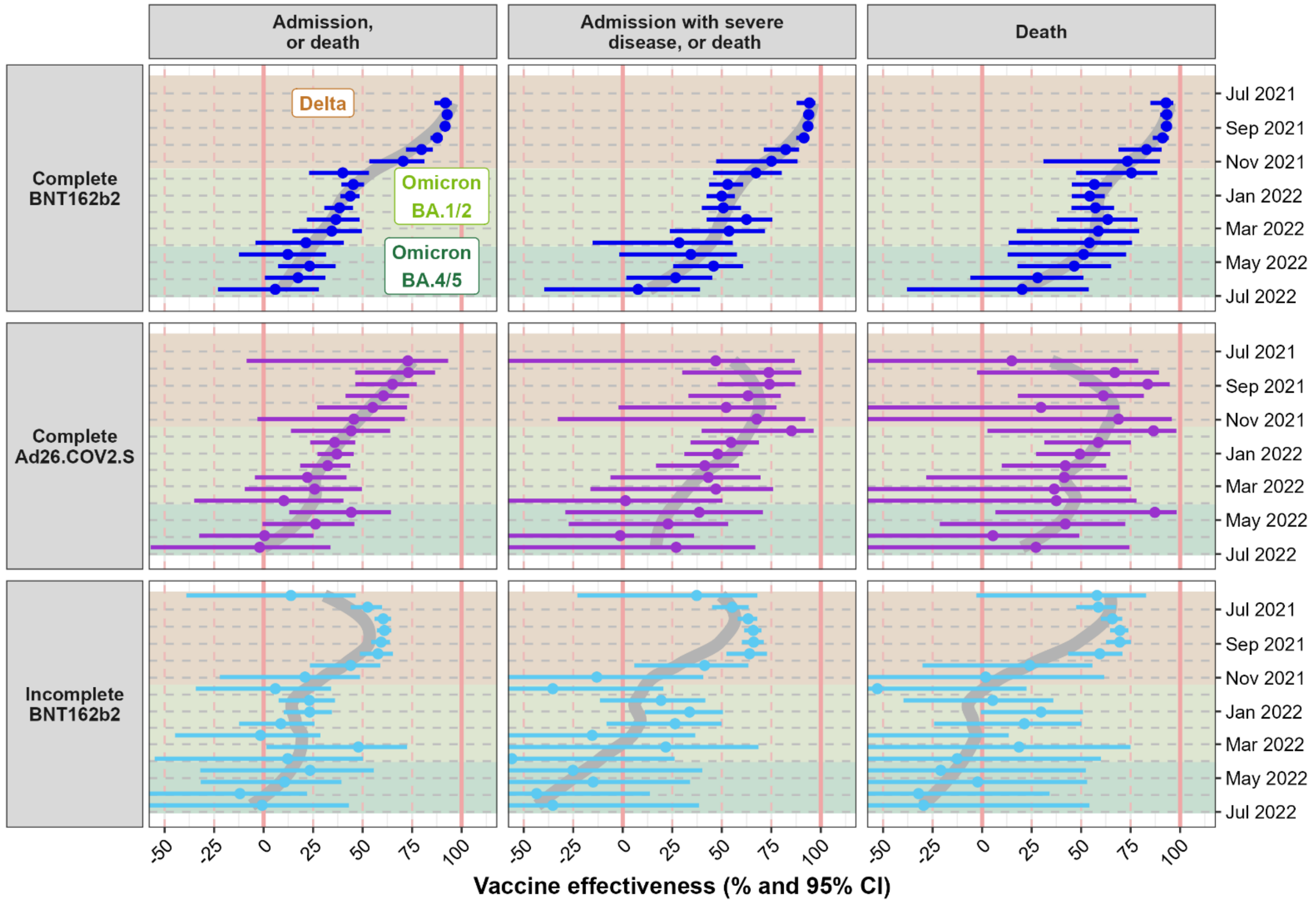
