## Appendix A for "COVID-19 vaccine uptake and effectiveness by time since vaccination in the Western Cape province, South Africa: An observational cohort study during 2020-2022"

This appendix provides supplementary content about methods for the following article:

2024

### Table of contents

**A1. Definition of wave periods** **3**

**A2. Definitions of outcomes** **4**

**A3. Definition of vaccine states and time in vaccine state** **5**

A3.1 Vaccine states and time in vaccine state: Ad26.COV2.S 5

A3.2 Vaccine states and time in vaccine state: BNT162b2 6

**A4. Definition of cox model covariates** **7**

**A5. The effect of incomplete data on prior SARS-CoV-2 infection** **10**

A5.1 Definitions and notations for our model world 10

A5.2 Bias measures 11

A5.3 Results for a few scenarios 11

### A1. Definition of wave periods

In South Africa, there have been five distinct COVID-19 waves to date, during which different SARS-CoV-2 (sub)lineages were most dominant. To define the ‘wave’ analysis periods used in this study, for each (set of) (sub)lineage(s) that emerged as dominant at the start of each of the waves, we defined the start of the period as (the start of) the earliest epidemiological week during which the (sub)lineage(s) comprised at least 20% of sequenced specimens nationally, using data from the Network for Genomics Surveillance in South Africa[1] (total number of sequenced and assigned specimens until end 2022: 11 864). For each period, the prevalence of the dominant (sub)lineage(s) increased rapidly to above 50% within 1-2 weeks, and a surge in SARS-CoV-2 diagnoses was observed. For our analyses we considered each wave period to extend to the start of the next wave period.

The first wave began on 1 March 2020; and we defined the last (fifth) wave as ending when the count of weekly SARS-CoV-2 diagnoses in our cohort dropped below 50 for the first time since the start of the South African pandemic.

The dates of the wave periods follow:

| **Wave** | **Dominant (sub)lineage(s)** | **Vaccine availability** | **Analysis period** | |
|---|---|---|---|---|
| | | | **Start date** | **End date** |
| 1 | Ancestral | No | 01 Mar 2020 | 17 Oct 2020 |
| 2 | Beta | No | 18 Oct 2020 | 29 May 2021 |
| 3 | Delta | Yes | 30 May 2021 | 13 Nov 2021 |
| 4 | Omicron BA.1 and BA.2 | Yes | 14 Nov 2021 | 02 Apr 2022 |
| 5 | Omicron BA.4 and BA.5 | Yes | 03 Apr 2022 | 02 July 2022 |
| None | | Yes | 03 July 2022 | 31 Dec 2022 |

[1] Msomi et al. A genomics network established to respond rapidly to public health threats in South Africa. The Lancet Microbe 2020. 1(6): e229 - e230. Also see the SARS-CoV-2 surveillance updates at: https://www.nicd.ac.za/diseases-a-z-index/disease-index-covid-19/sars-cov-2-genomic-surveillance-update

### A2. Definitions of outcomes

| | Outcome | Description | Algorithm to identify outcome |
|---|---|---|---|
| 1 | **Hospitalisation, or death** | A SARS-CoV-2 diagnosis that was associated with a hospital admission, or that was associated with death (see definition below) | • A hospital admission date within 7 days before to 21 days after the date of the SARS-CoV-2 diagnosis; AND • The admission was not at a specialised facility (psychiatric or rehabilitation); AND • No occurrence of a death within 28 days after the admission date where the death was recorded as not natural in the vital registry OR • A COVID-19 related death outcome (see below) |
| 2 | **Hospitalisation with severe disease, or death** | A SARS-CoV-2 diagnosis that was associated with a hospital admission that required admission to the intensive care unit or the prescription of oral or intravenous steroids, or that was associated with death (see definition below) | • A COVID-19 related hospitalisation as per definition above; AND • Admission to the intensive care unit at any point during the hospitalisation OR the prescription of oral or intravenous steroids during the hospitalisation OR • A COVID-19 related death outcome (see below) |
| 3 | **Death** | A SARS-CoV-2 diagnosis that was associated with death | • A death date within 7 days before to 28 days after the date of the SARS-CoV-2 diagnosis; OR • A death date within both (i) 7 days before to 60 days after the date of the SARS-CoV-2 diagnosis, and (ii) 14 days of a hospital discharge date AND • No indication of a non-natural death as per the vital registry |

- The date of the outcome is set to the date of the SARS-CoV-2 diagnosis, which is the date of the reporting of the positive test result to the Western Cape Provincial Health Data Centre (PHDC).
- We did not consider as an outcome simply having a confirmed SARS-CoV-2 infection. This is because only a small percent of persons who experienced SARS-CoV-2 infections were tested and diagnosed[2], and therefore the identification of infections is driven by testing behaviour and severity of disease. More severe COVID-19 outcomes would be less influenced by testing behaviour as people would need to seek care due to disease symptoms and would be tested if symptom severity required hospital admission.

[2] Hussey H, Vreede H, Davies M-A, et al. Epidemiology and outcomes of SARS-CoV-2 infection associated with anti-nucleocapsid seropositivity in Cape Town, South Africa. medRxiv 2022:2022.12.01.22282927.

### A3. Definition of vaccine states and time in vaccine state

The figures below describe how the vaccine terms used in the analysis (‘vaccine state’ and ‘time in state’) were defined, and the movement of persons among categories, based on the type of the first vaccine – either Ad26.COV2.S or BNT162b2.

#### *A3.1 Vaccine states and time in vaccine state: Ad26.COV2.S*

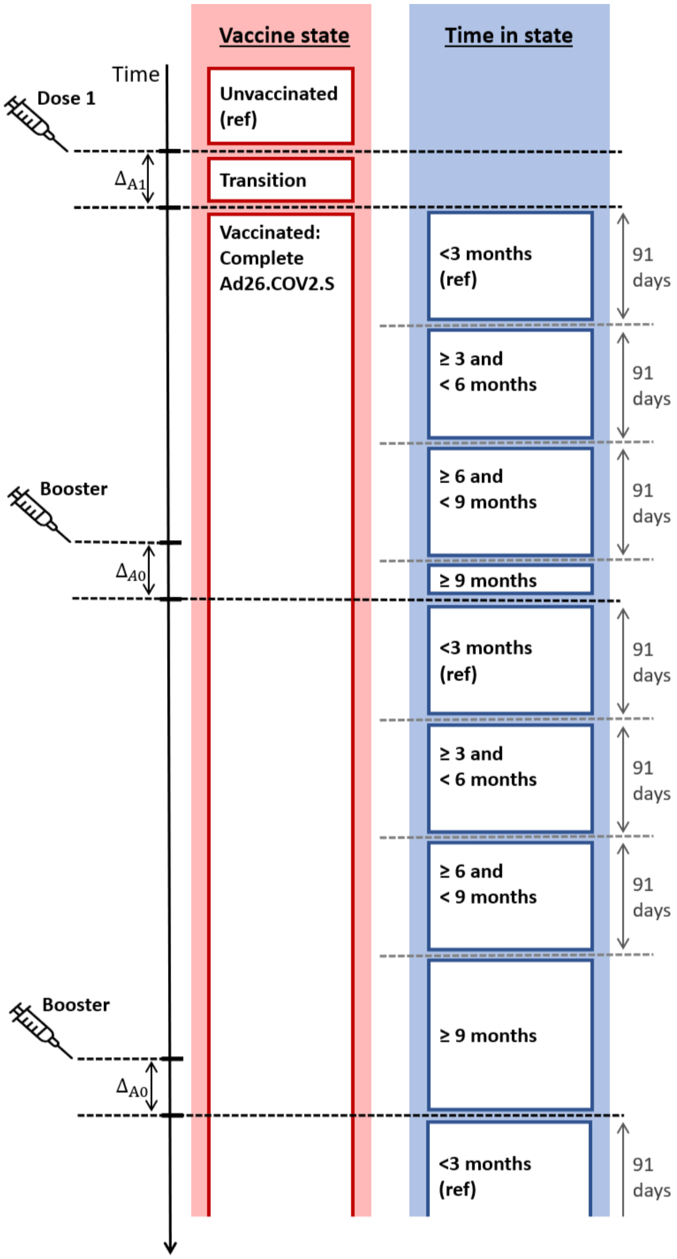


where

- $\Delta_{A1}$ = 28 days
- $\Delta_{A0}$ = 7 days

#### *A3.2 Vaccine states and time in vaccine state: BNT162b2*

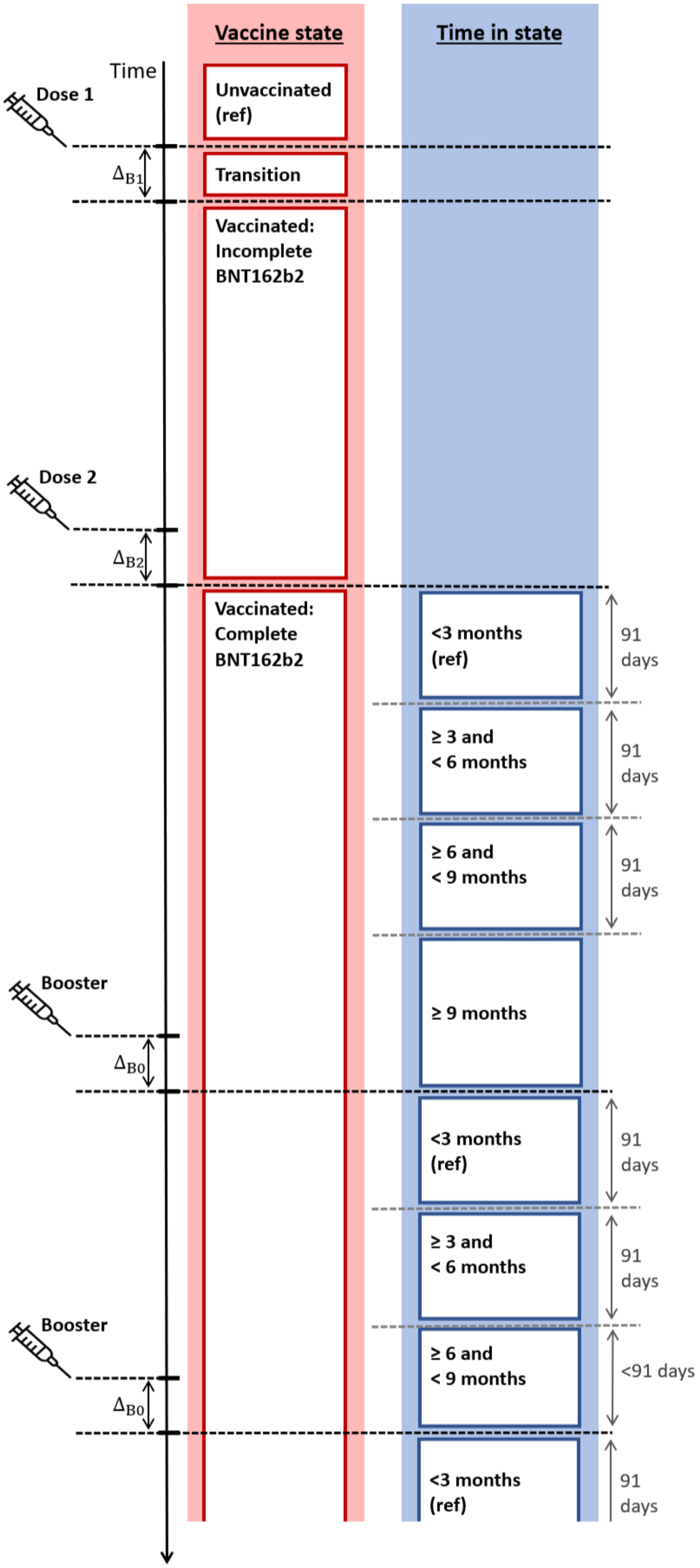


where

- $\Delta_{B1}$ = 21 days
- $\Delta_{B2}$ = 14 days
- $\Delta_{B0}$ = 7 days

Also, for a vaccine to be considered the second dose in the primary vaccination series, it had to be within 6 months of the previous dose, though an additional month was allowed if there was a recorded SARS-CoV-2 diagnosis between the doses as people were advised to delay receiving a vaccine after an infection. If the dose was not within this period, the person remained in the Incomplete BNT162b2 state, until the conditions were met to move into the Complete BNT162b2 state.

### A4. Definition of cox model covariates

| | | Covariate | Time-varying per person | Description and categories |
|---|---|---|---|---|
| **Demographic characteristics** | **1** | **Sex** | No | • Binary indicator: male versus female (reference) |
| | **2** | **Age** | Yes | • Age at current analysis calendar time<br>• Ordinal categories (in years): 18-34, 35-49 (reference), 50-59, 60-69, 70-79, ≥80 |
| **Non-communicable conditions** | | **Hypertension** | Yes | • Binary indicator: present versus absent (reference).<br>• A person transitions into the state of having the condition 90 days before the first recorded evidence, and remains in the state |
| | | **Diabetes** | Yes | • Binary indicator: present versus absent (reference)<br>• A person transitions into the state of having the condition 90 days before the first recorded evidence, and remains in the state |
| | | **Chronic kidney disease (CKD)** | Yes | • Binary indicator: present versus absent (reference)<br>• A person transitions into the state of having the condition 90 days before the first recorded evidence, and remains in the state |
| | | **Chronic respiratory disease** | Yes | • Binary indicator: present versus absent (reference)<br>• This condition includes both chronic obstructive pulmonary disease (COPD) and asthma (which we are unable to differentiate as medication overlaps)<br>• A person transitions into the state of having the condition, and remains in the state, 90 days before the first recorded evidence; unless there is a COVID-19 diagnosis date close to the first evidence date. If there is COVID-19 diagnosis within 14 days after or 90 days before the first recorded chronic respiratory disease evidence, the person enters the state 90 days after the latest COVID-19 diagnosis meeting this criterion[3]. Note that by definition a person is not at risk of another COVID-19 diagnosis for 90 days after a previous COVID-19 diagnosis, and does not contribute time at risk to the analyses of outcome rates during these 90 days. |

[3] This is because evidence of chronic respiratory disease (prescription of inhaled steroids or bronchodilators) could be directly related to the COVID-19 episode rather than due to pre-existing chronic respiratory disease, and therefore we only want to consider the person as already having the risk factor of chronic respiratory disease (which could impact subsequent COVID-19 outcomes) after the current COVID-19 episode.

| | | Covariate | Time-varying per person | Description and categories |
|---|---|---|---|---|
| **Infectious conditions** | | **HIV** | Yes | • Binary indicator: present versus absent (reference)<br>• A person transitions into the state of having the condition 90 days before the first recorded evidence, and remains in the state |
| | | **Any (ongoing/ previous) tuberculosis** | Yes | • Binary indicator: present versus absent (reference)<br>• A person transitions into the state of having the condition 30 days before the first recorded evidence of tuberculosis, and remains in the state<br>• Note that a person can be in this state due to an ongoing episode, or a previously experienced one |
| | | **An ongoing episode of tuberculosis** | Yes | • Binary indicator: present versus absent (reference)<br>• For each distinct inferred tuberculosis episode during the analysis period, the person transitions into the state of having the condition 30 days before the first recorded evidence for the episode of tuberculosis, and exits the state 9 months later<br>• In the multivariable cox model, the effect for this term is, among people with any tuberculosis (see covariate definition in row above), the *additional* effect of having an ongoing episode, rather than only one in the past |
| **Other conditions** | | **Pregnancy** | Yes | • Binary indicator: present versus absent (reference)<br>• For each pregnancy, a person transitions into the state on the estimated pregnancy start date and exits on the estimated pregnancy end date |
| **Prior SARS-CoV-2 diagnosis** | | **Prior SARS-CoV-2 diagnosis** | Yes | • Binary indicator: present versus absent (reference)<br>• Immediately after a SARS-CoV-2 diagnosis date, the person enters this state and remains in it |
| **Location characteristics** | **3** | **PHC visit location type** | No | • Binary indicator of the type of location where the person last accessed a primary healthcare facility, before the COVID-19 pandemic: Cape Town Metro versus other (reference), where other is non-Metro or no recorded visit. |
| | **4** | **District/ subdistrict** | No | • Geographical area (nominal categories), specified as the subdistrict or district where the person last accessed a primary care facility[4].<br>• For the City of Cape Town Metropolitan Municipality (the largest district in the Western Cape, containing 61% of our study population, and constituting eight subdistricts), the location is specified at a subdistrict level.<br>• For the remaining five districts, the location is specified at a district level. |

[4] Or, if not available, had a last encounter with a public sector healthcare facility, or, if also not available, last had an admission.

| | | Covariate | Time-varying per person | Description and categories |
|---|---|---|---|---|
| **Healthcare utilisation summary measures** | | **Number of previous negative tests** | No | • Ordinal categories (number of tests): 0 (reference), 1, ≥2<br>• Individual tests were first collapsed into distinct 'testing events'. All tests that were ≤2 days after a previous test were part of the same testing event. If any of the tests classified as belonging to the same testing event were positive, then the overall result was positive, and the testing event would not lead to an increment in the number of negative tests. If they were all negative, then the overall result was negative, and the number of previous negative tests would increase by 1, on the day of the first test of that testing event.<br>• For each analysis, over a selected period of calendar time, this counts previous negative tests from the start of the pandemic (March 2020) until 90 days before the start of the calendar period analysed. |
| | | **Number of visits at a primary healthcare facility** | No | • Ordinal categories (number of visits): 0 (reference), 1-4,5-14, ≥15<br>• The number of recorded visits at public healthcare primary care facilities in the 5 years before the pandemic (March 2020). |
| | | **Number of years with primary healthcare facility visits** | No | • Ordinal categories (number of years): 0 (reference), 1, 2, 3<br>• A count of the number of years (out of 3) with at least one recorded public healthcare primary care facility visit, during the three years before the pandemic (March 2020). |

### A5. The effect of incomplete data on prior SARS-CoV-2 infection

In South Africa, the ascertainment of SARS-CoV-2 infections is very low because of limited testing. The impact of incomplete data on the 'prior SARS-CoV-2 infection' status of a person on 'vaccine effectiveness' estimation is explored in a simple model world below. In this model world, each person has two binary characteristics: unvaccinated or vaccinated, which is recorded in the data; and has had a prior infection or not, which is not recorded in the data. Each person may experience a defined COVID-19 outcome.

#### *A5.1 Definitions and notations for our model world*

**Vaccine status** is denoted by subscript $i \in \{U, V\}$, where $U$ represents being unvaccinated, and $V$ being vaccinated.

**Prior infection status** is denoted by subscript $j \in \{P, S\}$, where $P$ represents having experienced a prior SARS-CoV-2 infection, and $S$ having not done so (i.e., still being susceptible to a first infection).

In our population, over a short study period:

- $\lambda_0$ = the **rate of the outcome** in unvaccinated (U) susceptible (S) persons
- $\theta_{V|j}$ = the direct (multiplicative) **impact of being vaccinated** (versus being unvaccinated) on the rate of the outcome, among persons with prior infection status $j$.
- $\theta_{P|i}$ = the direct (multiplicative) **impact of prior infection** (versus being susceptible) on the rate of the outcome, among persons with vaccine status $i$.
- $P_i$ = among persons with vaccination status $i$, the proportion of people with prior infection. i.e. the **prevalence of prior infection**.

Ideally, of interest would be the measurement of:

- $\tau_{V|S} = 1 - \theta_{V|S}$, the **true vaccine effectiveness in people without a prior infection**
- $\tau_{V|P} = 1 - \theta_{V|P}$, the **true vaccine effectiveness in people with prior infection**

However, in the absence of reliable data on the prior infection status of persons, in an analysis that does not include prior infection data, we instead can only measure some context-dependent association of vaccination and the outcome, $\tau = 1 - \theta$, where

$$\begin{aligned}
\theta &= \frac{\text{Outcome rate in vaccinated}}{\text{Outcome rate in unvaccinated}} \\
&= \frac{P_V \cdot \lambda_0 \cdot \theta_{V|P} \cdot \theta_{P|V} + (1 - P_V) \cdot \lambda_0 \cdot \theta_{V|S}}{P_U \cdot \lambda_0 \cdot \theta_{P|U} + (1 - P_U) \cdot \lambda_0} \\
&= \frac{P_V \cdot \theta_{V|P} \cdot \theta_{P|V} + (1 - P_V) \cdot \theta_{V|S}}{P_U \cdot \theta_{P|U} + (1 - P_U)}
\end{aligned}$$

Below we **compare values of the 'vaccine effectiveness' we can measure, $\tau$, to the true input values of vaccine effectiveness $\tau_{V|P}$ (in those with prior infection) and $\tau_{V|S}$ (in those without)** for a few different scenarios.

#### A5.2 Bias measures

The plots below show how (i) the ‘vaccine effectiveness’ quantity we can measure (left subplot), and (ii) bias in the estimation of vaccine effectiveness (middle and right plot), vary as a function of the prevalence of prior infection in vaccinated (x-axis) and unvaccinated (y-axis) persons.

Bias is defined as the relative error in our measurement (expressed as a percent), comparing the association that we can measure to: (i) the true vaccine effectiveness in the susceptible population (middle plot), or (ii) the true vaccine effectiveness in the prior-infected population (right plot).

Expressing the plotted quantities using the notation above:

- The left plot (‘Measurable vaccine effectiveness’) reports the association we can measure $\tau = 1 - \theta$
- The middle plot (‘Bias (%): susceptible population’) reports the bias $\frac{\tau - \tau_{V|S}}{\tau_{V|S}}$
- The right plot (‘Bias (%): prior-infected population’) reports the bias $\frac{\tau - \tau_{V|P}}{\tau_{V|P}}$

#### A5.3 Results for a few scenarios

##### No impact of prior infection on outcomes, equal vaccine effectiveness in prior-infected and susceptible

In this scenario, there is no bias in estimation of vaccine effectiveness.

##### No impact of prior infection on outcomes, different vaccine effectiveness in prior-infected and susceptible

In this scenario, the measurable association is some average of the vaccine effectiveness in the susceptible population and prior-infected population, and the weighting in this average will depend only on the prevalence of prior infection in the vaccinated population. The bias for an example scenario is shown below.

Scenario: Vaccine effectiveness in susceptible and prior-infected persons of 90% and 50% respectively.

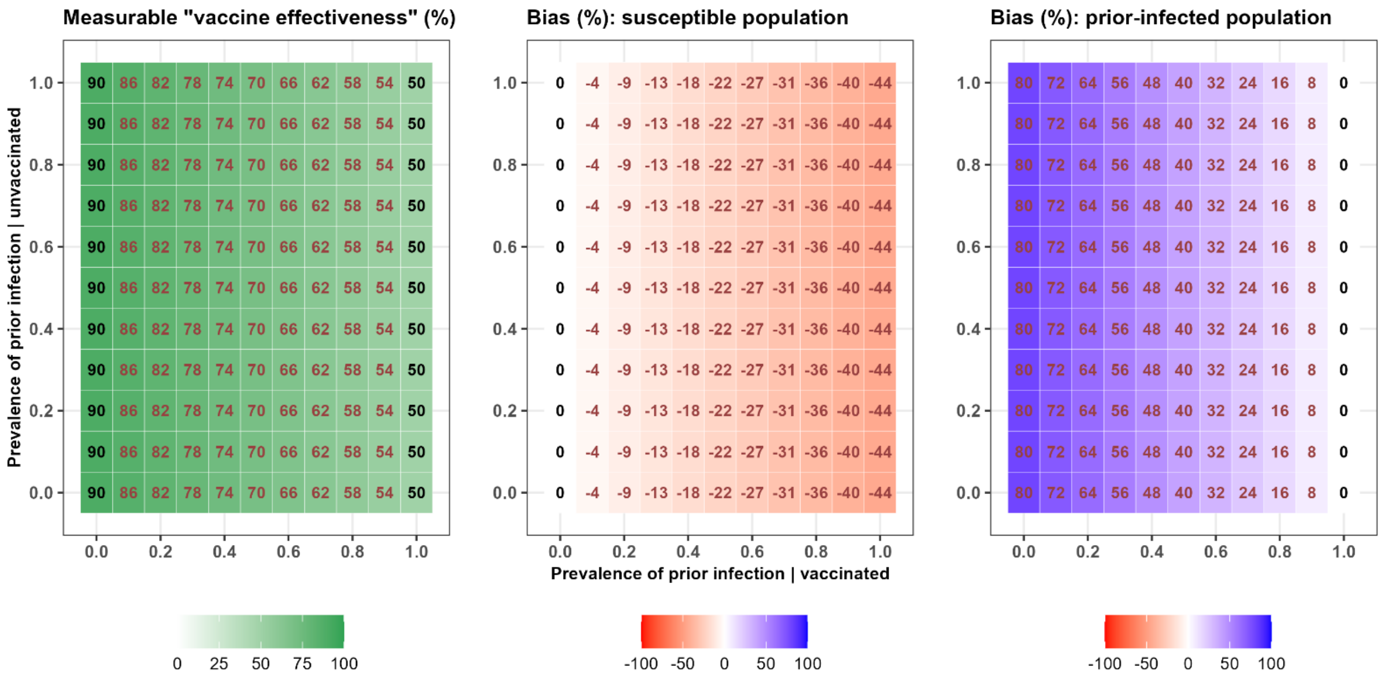


#### *Some impact of prior infection on outcomes*

In this setting, any difference in the prevalence of prior infection levels in the vaccinated and unvaccinated means that the two groups are already not comparable in terms of outcome rates, and biases will occur.

When there is a difference in vaccine effectiveness in prior-infected and susceptible persons, there is also an averaging of effects, resulting in a bias even when there is no difference in the prevalence of prior infection (in vaccinated versus unvaccinated persons). When there is a different impact of prior infection depending on vaccination status, again there will be a bias even when there is no difference in prevalence.

See example scenarios below.

Scenario: Vaccine effectiveness in susceptible and prior-infected persons of 70% in each group, and a reduction in outcome rate for persons with prior infection compared to susceptible persons of 70% (regardless of vaccine status)

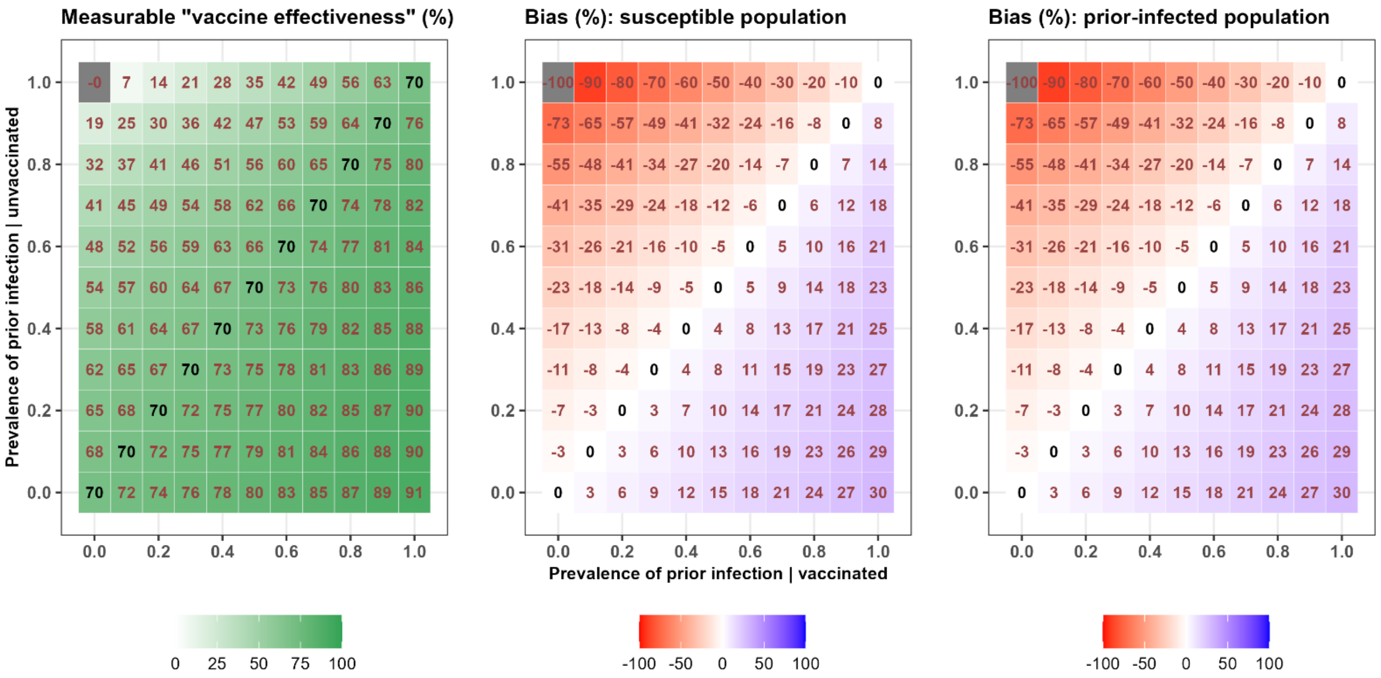


Scenario: Vaccine effectiveness in susceptible and prior-infected persons of 90% and 50% respectively, and a reduction in outcome rate for persons with prior infection compared to susceptible persons of 70% (regardless of vaccine state)

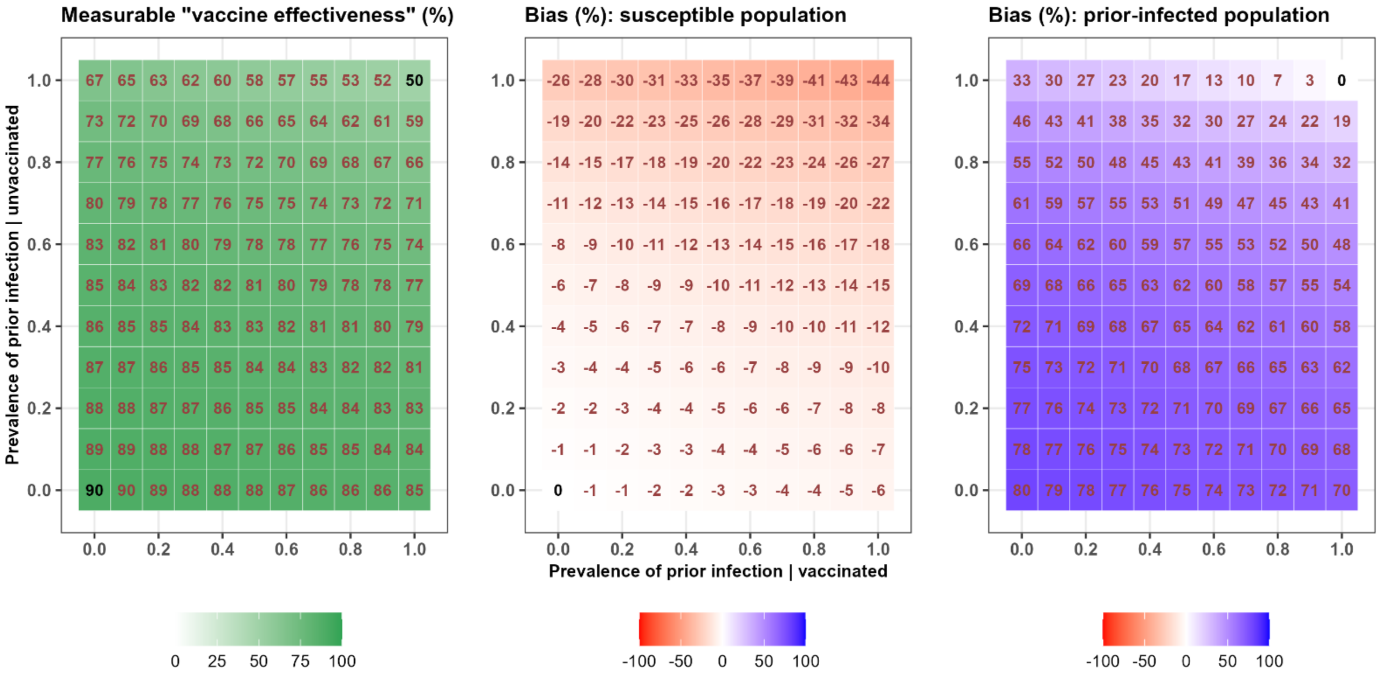


13 **Appendix A: Methods**
See Kassanjee et al. COVID-19 vaccine uptake and effectiveness by time since vaccination in the Western Cape province, South Africa: An observational cohort study during 2020-2022.

Scenario: Vaccine effectiveness in susceptible and prior-infected persons of 70% in each group, and a reduction in outcome rate for persons with prior infection compared to susceptible persons of 90% and 50% for unvaccinated and vaccinated persons respectively.

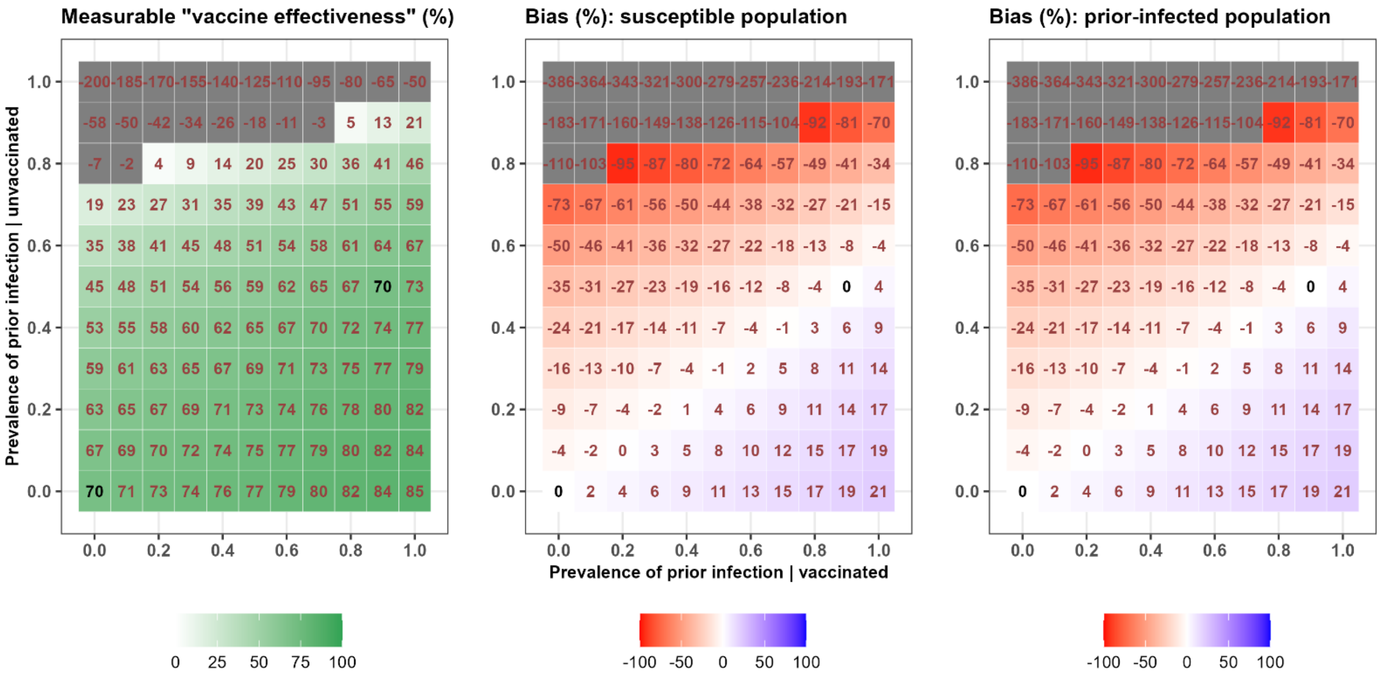


Scenario: Vaccine effectiveness in susceptible and prior-infected persons of 90% and 50% respectively, and a reduction in outcome rates for persons with prior infection compared to susceptible persons of 90% and 50% for unvaccinated and vaccinated persons respectively.

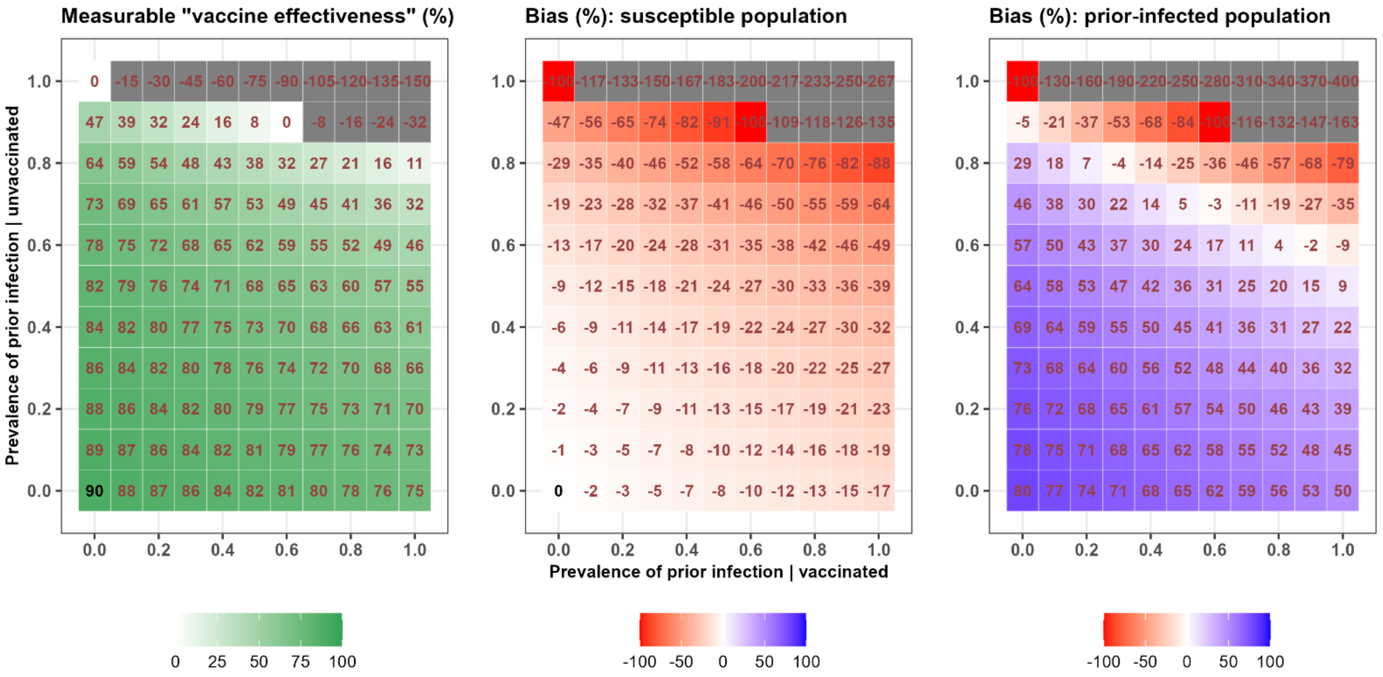



**Appendix A: Methods**
See Kassanjee et al. COVID-19 vaccine uptake and effectiveness by time since vaccination in the Western Cape province, South Africa: An observational cohort study during 2020-2022.
